# Bayesian Learning to Reduce Cardiac Risk for Locally Advanced NSCLC Patients Based on Personalized Radiotherapy Prescription

**DOI:** 10.1101/2025.09.15.25335723

**Authors:** Ruitao Lin, Mei Chen, Xiaodong Zhang, Tianlin Xu, Ting Xu, Rachel C Maguire, Kelsey L Corrigan, Efstratios Koutroumpakis, Joe Y Chang, Steven H Lin, Aileen B Chen, Quynh-Nhu Nguyen, Saumil J Gandhi, Matthew S Ning, Julianna Bronk, Stephen Chun, Ali Ajdari, Joshua S Niedzielski, Jinzhong Yang, Xinru Chen, Tinsu Pan, Qing H Meng, Anne S Tsao, Anita Deswal, Radhe Mohan, Zhongxing Liao

**Affiliations:** Department of Biostatistics, Division of Discovery Science, The University of Texas MD Anderson Cancer Center, Houston, Texas, USA; Department of Thoracic Radiation Oncology, Division of Radiation Oncology, The University of Texas MD Anderson Cancer Center, Houston, Texas, USA; Department of Radiation Physics, Division of Radiation Oncology, The University of Texas MD Anderson Cancer Center, Houston, Texas, USA; Edwards Lifesciences, Irvine, CA, USA; Department of Radiation Oncology, Division of Radiation Oncology, The University of Texas MD Anderson Cancer Center, Houston, Texas, USA; Department of Cardiology, Division of Internal Medicine, The University of Texas MD Anderson Cancer Center, Houston, Texas, USA; Department of Radiation Oncology, Massachusetts General Hospital and Harvard Medical School, Boston, Massachusetts, USA; The University of Texas MD Anderson Cancer Center UTHealth Houston Graduate School of Biomedical Sciences, Houston, Texas, USA; Department of Imaging Physics, Division of Diagnostic Imaging, The University of Texas MD Anderson Cancer Center, Houston, Texas, USA; Department of Laboratory Medicine, Division of Pathology/Lab Medicine, The University of Texas MD Anderson Cancer Center, Houston, Texas, USA; Department of Thoracic/Head and Neck Medical Oncology, Division of Cancer Medicine, The University of Texas MD Anderson Cancer Center, Houston, Texas, USA

## Abstract

**Purpose:** Radiation-induced heart damage is a significant concern in the treatment of non-small cell lung cancer (NSCLC) that can have debilitating or life-threatening consequences. Current strategies focus on minimizing heart exposure, but individual susceptibility varies. Existing evidence also suggests that a uniform “one-size-fits-all” dosimetric constraint for the heart may not be optimal for all patients.

**Methods:** We developed a prospective study using Bayesian continuous learning and adaptation to develop a framework for personalized adaptive radiation treatment (PART) to reduce cardiovascular adverse events (CAEs) among patients with locally advanced NSCLC. The trial includes a Bayesian personalized risk prediction model to guide heart dose constraints; sequential learning to refine the model and the PART; continuous adaptation of the target risk level; and go/no-go monitoring of PART effectiveness in clinical implementation. Elevation of high-sensitivity cardiac troponin T (hs-cTnT) after radiation was used as a surrogate biomarker for grade ≥2 CAEs to allow real-time decision-making.

**Results:** As of July 31, 2025, 100 patients have been enrolled and completed radiation treatment. Standard radiation plans were implemented for cohort 1 (50 patients), and PART for cohort 2 (50 patients). The first model incorporated patient- and disease-related factors and mean heart dose (MHD) as risk factors. The average treated MHDs were 7.84 ± 6.30 Gy in cohort 1 and 6.36 ± 6.01 Gy in cohort 2. The incidence of hs-cTnT elevation was lower in cohort 2 (20.5%) than in cohort 1 (31.9%). Within cohort 2, patients who satisfied the PART dose constraint had a markedly lower incidence of hs-cTnT elevation (9.7%) compared with those who exceeded the PART dose constraint (46.2%)..

**Conclusion:** Clinical implementation of PART model to guide treatment decision within a prospective trial is feasible. The recommended mean heart dose constraints generated by the first version of PART appear reasonable and clinically relevant. PART was associated with lower incidence of hs-cTnT elevation.

## INTRODUCTION

Undergoing radiation therapy for non-small cell lung cancer (NSCLC) carries the risk of radiation-induced adverse events from inadvertent exposure of organs at risk, with the risk depending on the radiation dose to specified volumes of those organs. Cardiac adverse events (CAEs) constitute a major category of treatment-induced toxicity after thoracic radiotherapy. Radiation-induced heart disease (RIHD) can have significant effects on survival and quality of life after treatment ^1–5^. The incidence of symptomatic cardiac events in NSCLC patients can be as high as 28.6%, and such events can occur as early as 3 months after chemoradiation ^6^. Unlike other types of treatment-induced toxicity, radiation-induced cardiac injuries are largely irreversible and can have debilitating and even life-threatening effects.

One potential strategy to limit radiation-induced injury is the use of advanced radiation modalities, such as volumetric modulated arc therapy (VMAT), passively scattered proton therapy (PSPT), or intensity-modulated proton therapy (IMPT), to minimize undesired radiation exposure to normal organs or their substructures. Proton therapy, in general, has been shown to spare at-risk organs owing to its compact dose deposition patterns and abrupt drop-off in dose at the distal edge. However, existing analyses in the general population suggest conflicting evidence regarding the association between cardiac toxicity and treatment modality or mean heart dose (MHD). For example, in our randomized trial comparing PSPT with intensity-modulated radiation therapy for locally advanced NSCLC^7^, we observed that while PSPT reduced MHD, it did not demonstrate a statistically significant reduction in cardiac toxicity (hazard ratio [HR]: 0.66, P = 0.306).^8^ We also found that changes in high-sensitivity cardiac troponin T (hs-cTnT) levels were highly associated with cardiac toxicity and strongly correlated with MHD.^8^ These findings in the general population suggest that susceptibility to cardiac toxicity varies among patients, indicating that a uniform “one-size-fits-all” MHD constraint may not be optimal for all patients. Instead, some patients may require stricter MHD controls, while others may not need such stringent constraints. To further complicate the issues, patients with lung cancer diagnosed at a median age of 65-67 years often present with pre-existing cardiac conditions, smoking-related comorbidities, diabetes, and others.

An optimal treatment intended to minimize RIHD risk may involve the following key steps. First, the risk of complications, such as RIHD, is estimated using an appropriate and robust prediction model. A personalized radiation plan is then developed to maintain the RIHD risk at a predefined level before radiation begins. Second, since RIHD typically develops late after the 6- to 7-week radiation course, an early endpoint enables timely monitoring and adaptation to keep RIHD incidence at or below the predefined level while ensuring the prediction model is updated appropriately. Third, the predefined RIHD level could be further refined based on information gained from the first two steps. These processes can be integrated into a prospective trial with flexible adaptation of treatment parameters (e.g., dose, duration, modality, or other medical intervention) based on continuous reassessment of previously enrolled patients. Such a trial would also allow for the prospective evaluation of treatment adaptations and their effectiveness ^9^ ^10^.

Recognizing the need for a prospective framework and workflow to allow risk to be predicted, reduced by treatment individualization, and adapted in a stepwise fashion, we initiated a prospective adaptive proof-of-concept study with the aim of developing a clinically applicable framework that incorporates personalized adaptive radiation treatment planning (PART). This study is guided by an individual risk model of CAEs and uses Bayesian machinery to guide the development of the PART. It also continuously monitors the effectiveness of the sequentially optimized PART. The objectives of this paper are to (1) introduce our Bayesian study design and the first-version PART model, (2) demonstrate the feasibility of co-developing and co-evaluating PART within a trial, and (3) present preliminary data from the first two patient cohorts.

## PATIENTS AND METHODS

### Trial objectives and endpoints

This trial enrolled patients aged ≥ 18 years with a histologic diagnosis of non-small cell lung cancer, who provided informed consent and for whom thoracic radiation therapy combined with concurrent systemic therapy was the recommended treatment. Informed consent is required for each patient before enrollment. A detailed description of the eligibility criteria can be found in the trial protocol. The following three objectives were derived through extensive deliberation among investigators, including radiation oncologists, cardiologists, radiation physicists, and biostatisticians:

1. We will develop a clinically relevant personalized normal tissue complication probability (NTCP) model for predicting risk of CAEs. This model will incorporate important clinical, biological, and dosimetric factors to forecast the probability of CAE and serve as a guide for developing PART, which will enable clinicians to prescribe radiation treatment doses and select radiation modalities tailored to individual profiles of cardiac toxicity risk.
2. We will explore the feasibility of using an optimized radiation modality and technique to achieve heart dose constraints to maintain the CAE risk at a preset NTCP level while still adhering to standard dosimetric constraints for the tumor and other organs at risk.
3. Throughout the trial, we will continuously monitor the effectiveness of the PART in reducing cardiac toxicity and quantify the reduction achieved.

The following co-primary endpoints were used in both model development and trial monitoring:

1. Elevation of hs-cTnT after radiation ^8^
2. Grade ≥2 CAEs within 12 months after radiation

Instead of directly using treatment-related cardiotoxicity (which may develop long after treatment) as the single primary endpoint, we also considered an early biomarker, the hs-cTnT level, as a co-primary endpoint. Use of an early biomarker can facilitate rapid decision-making and reduce the duration of a trial to a realistic timeframe and alleviate concerns associated with the unavailability of long-term cardiotoxicity data (i.e., missing data). Numerous studies have highlighted the ability of hs-cTnT assays to facilitate the early detection and prediction of cardiac toxicity induced by chemotherapy or radiotherapy^11,12^. Our previous multivariate analysis^8^ demonstrated that an increase of 5 ng/L during treatment (HR: 3.57, *p* = 0.009) was strongly predictive of CAE risk. In addition, the increase in hs-cTnT during treatment was significantly correlated with the mean heart dose (*p* = 0.0004). These findings highlight the potential of hs-cTnT as a dose-dependent biomarker for radiation-induced cardiac damage.

### Study design

We established a prospective Bayesian adaptive cohort study that incorporates two key Bayesian components: (1) a Bayesian risk prediction model for CAEs that accounts for patient-related and radiation-dosimetry parameters and (2) Bayesian sequential monitoring to evaluate the effectiveness of the developed PART in reducing the incidence of CAEs. Our goal was to enroll patients in sequential cohorts, each with a preset CAE target level, and use PART to progressively reduce the expected CAE risk in a stepwise manner, cohort by cohort.

We chose to develop and evaluate PART in a single-arm study using the first cohort as internal control for the following reasons. First, this is a proof-of-concept study, as we will learn and adapt our personalized radiation treatment plan as we go. Second, because PART modifies or reduces only the heart radiation dose, without compromising doses to tumors and other normal tissues, our intent to reduce toxicity without compromising treatment efficacy enhances the trial’s ethical standing. Third, our rich repository of historical trial data, along with data from patients treated off-protocol with standard-of-practice plans, will enable us to compare toxicity rates between PART and standard-of-practice treatments. Finally, having a single-arm trial aimed at reducing long-term heart toxicity would expedite trial enrollment.

The trial design (Figure 1) involves continuously conducting the following adaptations after each cohort of 50 patients is treated.

<u>(1) Personalized NTCP model adaptation</u> (“Model Adaptation” in Figure 1): After each cohort is treated, we will leverage the newly acquired response data, along with the data already acquired and additional risk factors (e.g., cardiac substructure dosimetric parameters, patient genetic radiosensitivity, use of systemic therapy) to refit the Bayesian personalized NTCP model for improvement.
<u>(2) Risk target adaptation</u> (“Target rate (X) adaptation” in Figure 1): Upon assessing the newly observed data, we will adaptively evaluate whether a lower cardiac toxicity risk level can be reasonably achieved based on our developed model.
<u>(3) Treatment adaptation</u> (Box “Treat with the PART plan” in Figure 1): We will adapt the radiation modality, technique, and dose distribution to examine if the recommended heart dose constraint can be achieved. If the desired dose constraint cannot be satisfied, we will do our best to reduce the high-risk dosimetric parameters for the patient.

**Figure 1.**
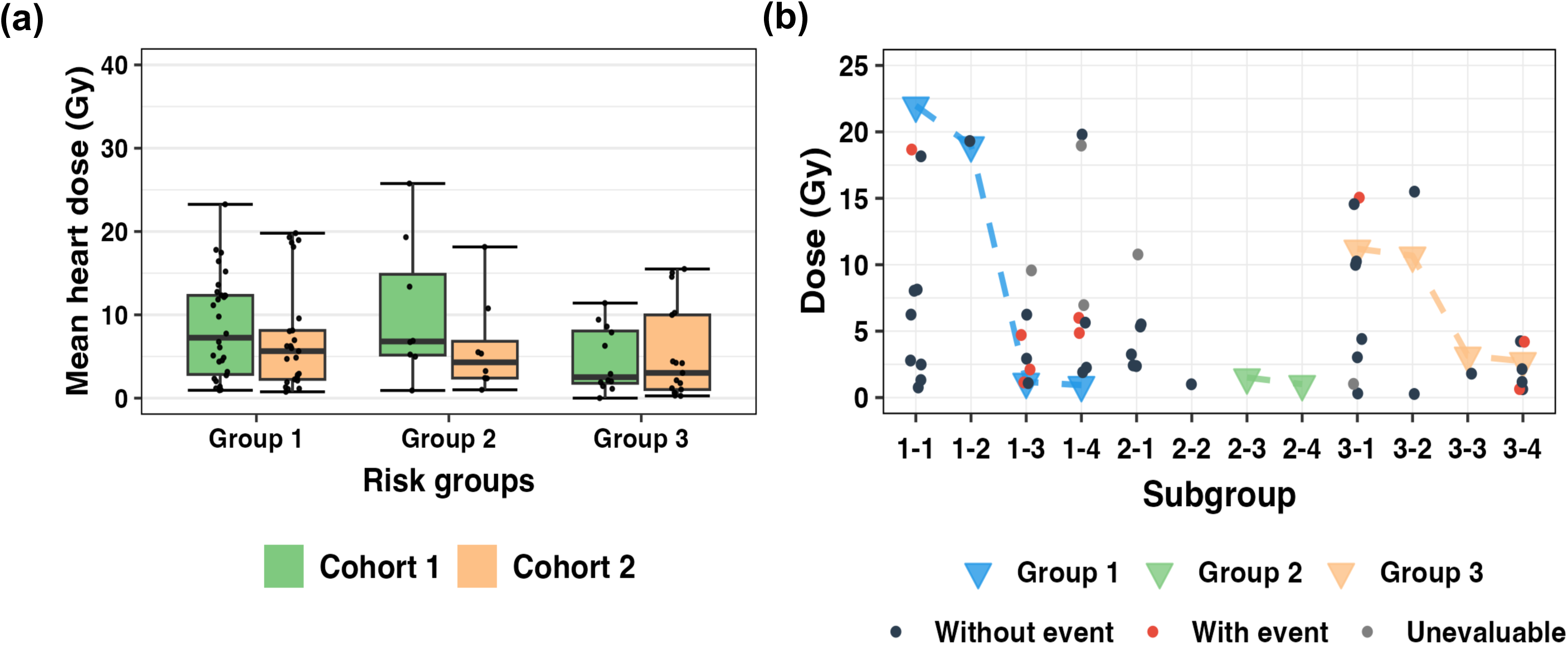
Schematic diagram of the adaptive treatment rule in this Bayesian adaptive cohort registration trial. After each cohort of patients is treated, if the toxicity rate is higher than the pre-specified boundary denoted as y, we will stop the patient enrollment; otherwise, enrollment continues, and our personalized normal tissue complication probability (NTCP) model is adaptively refined to derive the personalized radiation plan. The predicted risk using the standard plan is denoted by NTCP_S_, while the target risk level is denoted by X. If there is a high posterior likelihood that the cardiac risk induced by the standard radiation plan is greater than the pre-specified target X (denoted by Pr(NTCP_S_ > X | Data)), the patients should be treated with the personalized adaptive radiation treatment (PART) plan; otherwise, the patients should be treated with the standard plan. This process will be repeated until further reduction of CAE is not possible solely by radiation planning strategy, and at that time cardiac medical interventions for prevention or treatment will be introduced to mitigate cardiac adverse events.

We chose to use Bayesian machinery to adaptively develop the personalized radiation plan and monitor the added benefit in terms of risk reduction from the personalized plan, based mainly on the Bayesian’s core “learn-as-you-go” feature.^13^ Several other characteristics of the Bayesian approaches also contributed to the design:^14^ First, Bayesian methods can directly use the posterior probability that the toxicity rate of PART is well below a certain benchmark in decision-making. Second, Bayesian approaches can seamlessly incorporate prior or external information into the current study. Third, clinical implementation of a Bayesian hierarchical dose-response modeling approach models risk levels across different pre-defined patient subgroups so that information can be shared across different subgroups for improved estimation and decision-making.

### Personalized risk prediction model

We initially used MHD as the adaptable treatment variable to guide dose constraints in the radiation plan and reduce risk. By using predetermined variables, we formulated the following personalized NTCP model through a hierarchical logistic regression approach ^15^:

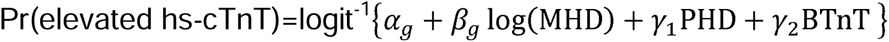

Here, PHD denotes the binary indicator for the pretreatment heart disease, BTnT is the baseline hs-cTnT level, the subscript *g* denotes the predefined risk groups (as detailed in the Results section). Within the Bayesian framework, prior distributions can be specified for the unknown parameters *α_g_*, *β_g_*, *γ_1_*, and *γ_2_*. The group-specific coefficients *α_g_* and *β_g_* can be estimated by leveraging information shared across these risk groups.

By deriving the posterior inference for the dose-toxicity curve for each specific patient subgroup, we then used these posterior quantities to assess whether the standard dosing plan is excessively toxic and, if so, to determine a safe dosing range based on the personalized NTCP model. Further details about the model, prior distribution, and two-step procedure are provided in the Data Supplement.

### Trial monitoring

To ensure that PART can effectively reduce treatment-related cardiac toxicity, we implemented the Bayesian optimal phase 2 (BOP2) design^16,17^ to makes adaptive “go/no-go” decisions^18^ by continuously monitoring the number of hs-cTnT elevations and 12-month treatment-related CAEs after treating each cohort of 50 patients with PART during the trial. If we find no significant reduction in the rates of hs-cTnT elevations and CAEs, relative to the historical rates from the use of standard radiation planning, we will stop future enrollment and conclude that the PART approach is not effective. This design has been shown to effectively control the false-positive rate while maximizing study power with limited sample sizes. Let *p*_1_ denote Pr(elevated hs-cTnT), and *p*_2_ denote Pr(grade ≥ 2 CAE). Based on an analysis of the completed trial of standard chemoradiation therapy for NSCLC ^8^, *p*_1_ was approximately 26%, and *p*_2_ was approximately 15% for standard planning. Under the null hypothesis, we would consider PART to be ineffective if *p*_1_ = 0.25 and *p*_2_ = 0.15, i.e., not reducing the historical rates. Under the alternative hypothesis, we would consider PART to be effective if *p*_1_ = 0.15 and *p*_2_ = 0.10, i.e., a decrease of 0.1 in Pr(elevated hs-cTnT) or a decrease of 0.05 in Pr(grade ≥ 2 CAE). Based on the details in the Data Supplement, the optimal decision boundaries for the BOP2 design are shown in Table S1 in the Data Supplement. Interim go/no-go decisions are made when the number of treated patients reaches 50, 100, and 150. At any point during the trial, enrollment will be stopped if the number of hs-cTnT elevations or 12-month CAEs exceeds the corresponding boundaries. When the trial reaches the maximum sample size of 200, PART will be considered effective if the number of hs-cTnT elevations is fewer than 38 or the number of grade ≥2 CAEs is fewer than 21.

## RESULTS

The protocol (ClinicalTrials.gov identifier: NCT05010109) was approved by our institutional review board on March 10, 2021, and the first patient was enrolled on August 12, 2021. The original design specified a cohort size of 25 patients per adaptation, with a total sample size of 100 patients. However, given the higher-than-anticipated accrual and the need for a larger sample size for model adaptation, a protocol amendment was made on June 23, 2023, increasing the cohort size to 50 and the total sample size to 200 patients. As of July 31, 2025, we have enrolled and treated 100 patients (50 in cohort 1 and 50 in cohort 2, Figure 2). The first cohort of patients were treated with standard heart dose constraints as a control group within the trial without PART. Consequently, the PART model was applied from cohort 2 onward.

**Figure 2.**
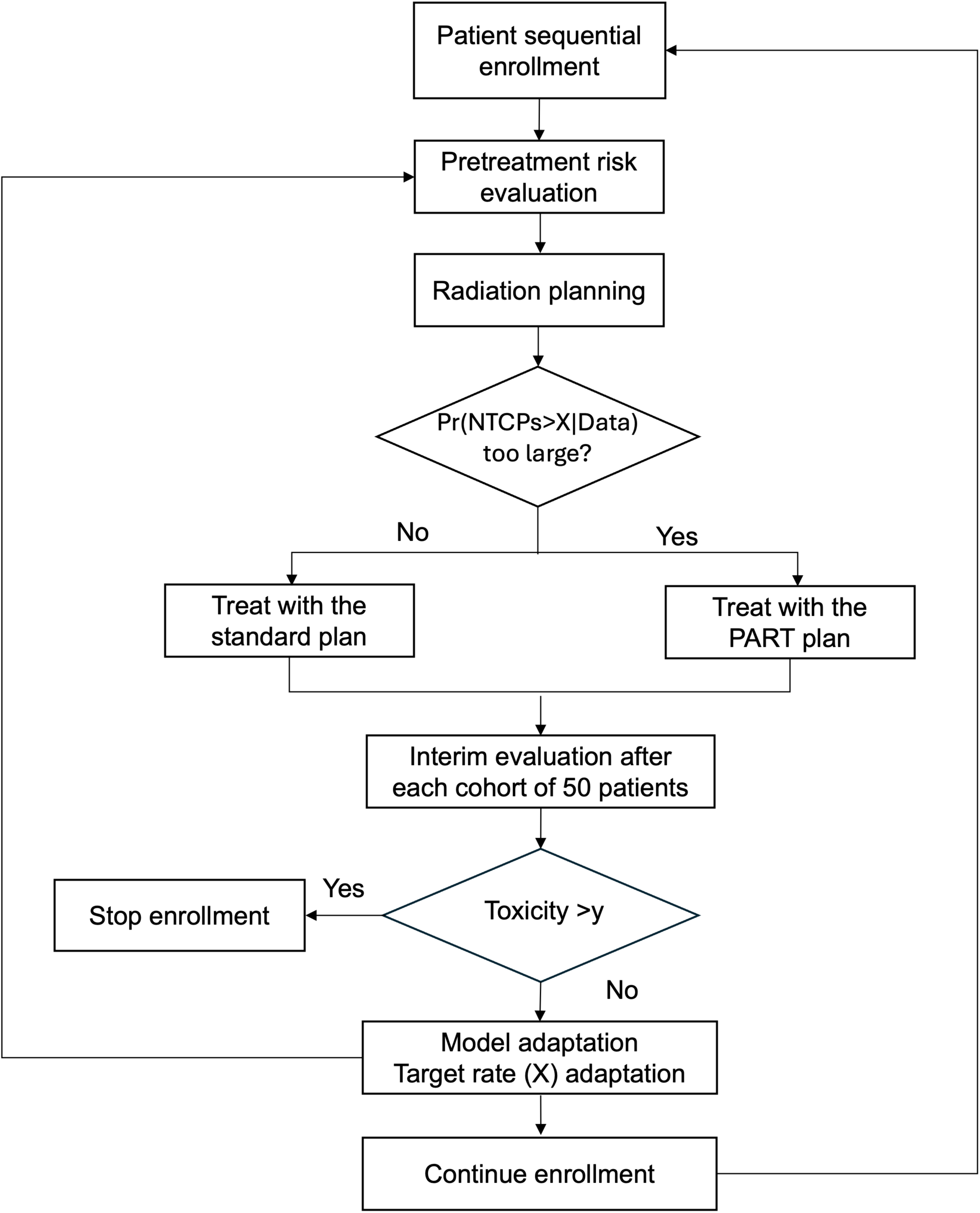
Overview of trial enrollment in the two cohorts as of July 31, 2025.

### Model development after cohort 1

Based on the data from our past randomized trial ^7^ and those of cohort 1, we identified three risk groups (Table 1) by using the classification tree approach. Within a group, the patients outcomes tended to be homogeneous and were associated with 18.9%, 16%, and 52.5% probabilities of elevated hs-cTnT. We also selected several other important risk factors and incorporated them into the personalized NTCP model, which included pre-existing heart disease and the baseline hs-cTnT status (whether it exceeded 10 ng/L or not). Because pretreatment hs-cTnT above 10 ng/L is associated with clinically symptomatic CAE (HR: 4.06, *p* = 0.005),^8^ and the extent of its elevation during chemoradiation therapy depended on MHD,^8^ MHD is currently used as a treatment parameter for dose constraints to keep the cardiac risk at or below the preset level.

**Table 1.** Summary statistics for the first two cohorts in this Bayesian adaptive cohort registration trial as of July 31, 2025.

|  |  | **Cohort 1** | **Cohort 2** | ***p*-value** |
| --- | --- | --- | --- | --- |
| **No. of enrolled patients** |  | 50 | 50 |  |
| **Age, y, mean ± SD** |  | 65.10 ± 9.14 | 68.42 ± 9.92 | 0.053 |
|  | Age≥64 | 29 (58%) | 36 (72%) | 0.208 |
|  | <64 | 21 (42%) | 14 (28%) |  |
| **Sex** |  |  |  | 0.543 |
|  | Male | 23 (46%) | 19 (38%) |  |
|  | Female | 27 (54%) | 31 (62%) |  |
| **Tumor location** |  |  |  | 0.421 |
|  | Right | 30 (60%) | 25 (50%) |  |
|  | Left/mediastinal | 20 (40%) | 25 (50%) |  |
| **Risk groups** |  |  |  | 0.518 |
|  | Group 1: Tumor location = right | 30 (60%) | 25 (50%) |  |
|  | Group 2: Tumor location = left/mediastinal, age <64 | 8 (16%) | 8 (16%) |  |
|  | Group 3: Tumor location = left/mediastinal, age ≥64 | 12 (24%) | 17 (34%) |  |
| **Pre-existing heart disease** |  |  |  | 1.000 |
|  | Yes | 18 (36%) | 19 (38%) |  |
|  | No | 32 (64%) | 31 (62%) |  |
| **Baseline hs-cTnT level, ng/L, mean ± SD** |  | 10.10 ± 8.66 | 16.39 ± 31.09 | 0.105 |
|  | >10 ng/L | 17 (34%) | 26 (52%) | 0.106 |
|  | ≤10 ng/L | 33 (66%) | 24 (48%) |  |
| **Treated mean heart dose, Gy, mean ±SD** |  | 7.84 ± 6.30 | 6.36 ± 6.01 | 0.202 |

Abbreviations: SD=standard deviation, hs-cTnT=high-sensitivity cardiac troponin T assay

This first simple model provided the recommended subgroup-specific MHD constraints of the initial PART. According to Table 2, if the goal is to reduce the risk levels of the three groups to 15%, 15%, and 45%, respectively, then for a patient with a tumor located on the right side, pre-existing heart disease, and a baseline hs-cTnT level below 10 ng/L, the recommended dose range is an MHD below 18.98 Gy. The maximum MHD under the target reduction of (15%, 15%, 45%) was adopted in the initial PART and implemented for cohort 2. An alternative approach could involve considering a more profound target risk reduction, such as (15%, 13%, 42%) or (13%, 11%, 37%). However, it is acknowledged that achieving the MHD range under this target would be challenging.

**Table 2.** Recommended maximum mean heart dose (Gy) based on the initial version of PART. The three numbers in the parentheses correspond to the target rates of cardiac event defined as elevated hs-cTnT greater than 5 ng/L in the three risk groups.

| Risk group | Pre-existing heart disease | Baseline hs-cTnT > 10ng/L | MHD to achieve risk reduction target |  |  |
| --- | --- | --- | --- | --- | --- |
|  |  |  | (15%, 15%, 45%) | (15%, 13%, 42%) | (13%, 11%, 37%) |
| 1<br>(Tumor location = right) | No | No | 21.97 | 21.97 | 18.57 |
|  | Yes | No | 18.98 | 18.98 | 18.98 |
|  | No | Yes | 1.18 | 1.18 | 1.51 |
|  | Yes | Yes | 0.94 | 0.94 | 1.18 |
| 2<br>(Tumor location = left/mediastinal, age <64) | No | No | No adaptation | 26.08 | 24.86 |
|  | Yes | No | No adaptation | 21.97 | 24.7 |
|  | No | Yes | 1.51 | 0.72 | 1.01 |
|  | Yes | Yes | 1.01 | 0.57 | 0.72 |
| 3<br>(Tumor location = left/mediastinal, age ≥64) | No | No | 11.20 | 9.49 | 8.32 |
|  | Yes | No | 10.68 | 9.11 | 8.47 |
|  | No | Yes | 3.15 | 2.74 | 2.74 |
|  | Yes | Yes | 2.74 | 2.19 | 2.19 |

Abbreviations: PART=personalized adaptive radiation treatment, hs-cTnT=high-sensitivity cardiac troponin T assay, MHD=mean heart dose

### Patient and treatment characteristics

The patients in cohort 1 and ongoing cohort 2 were similar in baseline characteristics, including age, sex, tumor location, pre-existing heart disease and baseline hs-cTnT level (Table 1). Cohort 2 patients were slightly older with a mean age of 68.42 ± 9.92 years compared to 65.10 ± 9.14 years in cohort 1, though this difference was not statistically significant (*p* = 0.053). Pre-existing heart disease prevalence was nearly identical between cohorts (36% vs 38%; *p* = 1.000). Baseline hs-cTnT levels showed a trend toward higher values in cohort 2 (cohort 2: 16.39 ± 31.09 ng/L, cohort 1: 10.10 ± 8.66 ng/L, *p* = 0.105), with more patients in cohort 2 having levels >10 ng/L (52% vs 34%, *p* = 0.106). As for the risk group stratification, the distribution was nearly balanced between cohorts with no statistically significant difference (*p* = 0.518).

PART evaluation was applied for each patient in cohort 2 (Figure 2). Of the 50 patients evaluated, the planned MHD of 34 patients (68%) met the dose constraint, and the MHD of 16 patients (32%) exceeded the dose constraint. The averaged MHDs were 4.41 ± 4.71 Gy and 9.65 ± 6.43 Gy for the groups met and exceeded the dose constraint, respectively. With the implementation of PART, the MHD in cohort 2 was 18.9% lower than that in cohort 1 (6.36 ± 6.01 Gy vs 7.84 ± 6.30 Gy, *p* = 0.202). Figure 3 (a) showed the MHD distribution of the three risk groups in both cohorts. Compared with cohort 1, lower median dose with less variability was observed in group 1 and group 2 of cohort 2. Group 3 showed the most similar distributions between cohorts.

**Figure 3.**
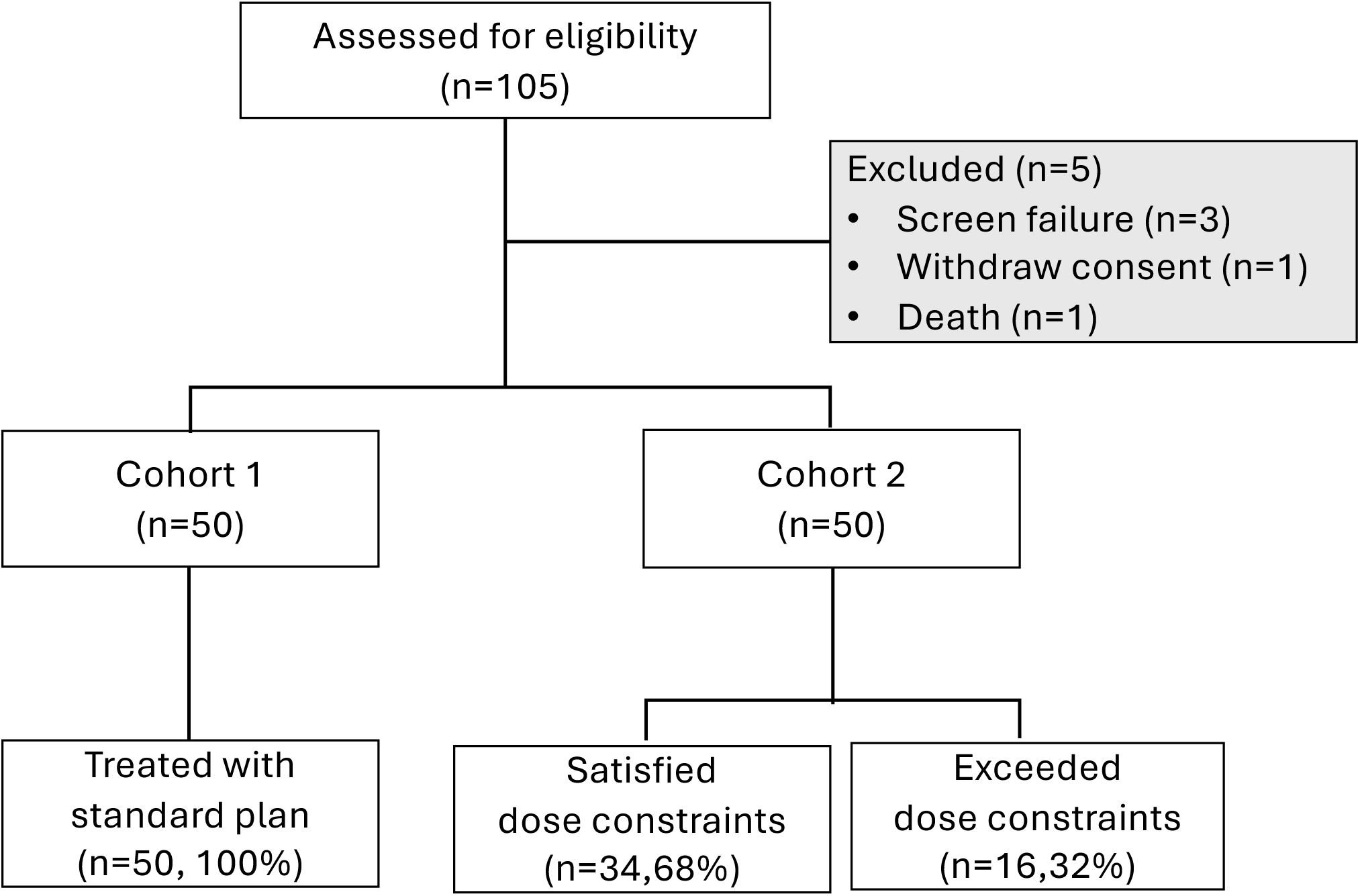
(a) Comparison of mean heart dose distribution in each risk group between cohort 1 (green) and cohort 2 (orange), the dose of each patient is shown in dot (b) personalized dose constraints (triangle) in each risk group and the mean heart dose of each patient (circle) in cohort 2, the event status was mark in red (with event), black (without event) and light gray (unevaluable).

### Elevation of hs-cTnT during radiation

As for the clinical outcome, only the hs-cTnT data during radiation are mature for analysis, whereas other clinical outcomes remain premature due to limited follow-up. Preliminary data (Table 3) showed that in cohort 1, the incidence of hs-cTnT elevation during radiation was 31.9% (15 of 47 evaluable patients, 95% CI: 0.19–0.47), while in cohort 2, the incidence was 20.5% (9 of 44 evaluable patients, 95% CI: 0.10–0.35). Based on unadjusted, inverse probability weighting (IPW)–adjusted, and regression-adjusted logistic regressions (details in the Supplementary Materials-1), the odds ratios comparing cohort 2 with cohort 1 ranged from 0.41 to 0.47, indicating a trend toward reduced hs-cTnT elevation in cohort 2.

**Table 3.** Comparison of hs-cTnT elevation rates between patient subgroups.

|  | Estimate | 95% CI | p-value |
| --- | --- | --- | --- |
| **(1) Comparing Patients in Cohort 1 and Cohort 2** |  |  |  |
| Event rate<sup>a</sup> |  |  |  |
| Patients in cohort 1 | 0.32 (15/47) | (0.19, 0.47) |  |
| Patients in cohort 2 | 0.20 (9/44) | (0.10, 0.35) |  |
| Average treatment effect<sup>b</sup> |  |  |  |
| Unadjusted | 0.47 | (0.19, 1.16) | 0.101 |
| IPW-adjusted | 0.41 | (0.16, 1.04) | 0.060 |
| Regression-adjusted | 0.46 | (0.17, 1.29) | 0.139 |
| **(2) Comparing PART and Non-PART Patients Within Cohort 2** |  |  |  |
| Event rate<sup>a</sup> |  |  |  |
| PART patients in cohort 2 | 0.10 (3/31) | (0.02, 0.26) |  |
| Non-PART patients in cohort 2 | 0.46 (6/13) | (0.19, 0.75) |  |
| Average treatment effect<sup>b</sup> |  |  |  |
| Unadjusted | 0.28 | (0.06, 1.18) | 0.083 |
| IPW-adjusted | 0.10 | (0.02, 0.62) | 0.013 |
| Regression-adjusted | 0.11 | (0.01, 0.84) | 0.033 |
| **(3) Comparing PART in Cohort 2 and Other Patients** |  |  |  |
| Event rate<sup>a</sup> |  |  |  |
| PART patients in cohort 2 | 0.10 (3/31) | (0.02, 0.26) |  |
| Other patients | 0.35 (21/60) | (0.23, 0.48) |  |
| Average treatment effect<sup>b</sup> |  |  |  |
| Unadjusted | 0.29 | (0.09, 0.86) | 0.026 |
| IPW-adjusted | 0.25 | (0.08, 0.82) | 0.022 |
| Regression-adjusted | 0.16 | (0.04, 0.65) | 0.010 |

Abbreviations: hs-cTnT=high-sensitivity cardiac troponin T assay, CI=confidence interval, IPW=Inverse probability weighting; PART = personalized adaptive radiation treatment
<sup>a</sup> Cases with missing hs-cTnT elevation data were excluded from calculating the event rate
<sup>b</sup> Logistic-regression models were used to estimate the odds ratio comparing subgroup 1 with subgroup 2. Missing data were handled using multiple imputation with 100 imputed datasets.

The relationship between planned MHD, personalized dose constraints, and event outcomes for individual patients in cohort 2 is visualized in Figure 3(b). The incidence of hs-cTnT elevation was 9.7% (3 of 31 evaluable patients, 95% CI: 0.02-0.26) among patients whose MHD met the dose constraints and 46.2% (6 of 13 evaluable patients, 95% CI: 0.19-0.75) among those who did not. To further evaluate the impact of meeting dose constraints with enhanced statistical power and control for confounding, we compared patients whose MHD satisfied the PART dose constraint (“PART patients”) with those who did not in cohort 2 (“Non-PART patients”), as well as with the combined group of all patients in cohort 1 together with Non-PART patients in cohort 2 (“Other patients”) under covariate adjustment. Table 3 demonstrates that after appropriate covariate adjustment (i.e., IPW-adjusted or regression-adjusted), meeting the PART dose constraint was associated with a statistically significant reduction in hs-cTnT elevation. For example, the estimated hs-cTnT elevation rate among PART patients in cohort 2 was 0.10 (95% CI: 0.02–0.26), compared with 0.35 (95% CI: 0.23–0.48) among other patients, with all odds ratio *p*-values < 0.05.

## DISCUSSION

We successfully developed a framework using a Bayesian adaptive sequential design to enable systematic continuous monitoring, learning, decision-making, and multi-level adaptations to improve patient outcomes, with encouraging preliminary results. Our current findings indicate that patients in this trial were older (median 68.5, IQR 61–73) and had high hs-cTnT levels at baseline (median 8.0 ng/L, IQR 3–14), compared to our previous randomized study^7,8^ (median age 66, IQR: 58–71; median baseline hs-cTnT level: 4.6 ng/L, IQR: 3.0–8.8). This trend aligns with reported cancer statistics, indicating that the median age of lung cancer patients is increasing and their survival is improving ^19^, despite older patients typically presenting with several cardiovascular-related comorbidities ^20,21^. In the original trial, we aimed to set a predefined CAE level with a progressive reduction goal for each cohort in a stepwise fashion. However, we encountered situations where the PART-recommended MHD was not achievable. Additionally, some patients required a change in radiation modality (e.g., proton therapy) to meet the PART MHD goal, but their insurance coverage was denied for that therapy. To address these logistic issues, we modified the cohort adaptation from a predefined goal to what PART could ultimately achieve, with the actual amount of reduction recorded and quantified for subsequent model adaptation. Encouragingly, the implementation of PART in cohort 2 achieved an 18.9% MHD reduction, lowering the dose from 7.84 ± 6.30 Gy to 6.36 ± 6.01 Gy. Moreover, reducing the MHD to meet the personalized dose constraint derived by the PART model was promising in reducing the incidence of hs-cTnT elevation during treatment, as evidenced by the lower incidence in the group that met the dose constraint (9.7%) than that in the group that exceeded the dose constraint (46.2%, *p* = 0.012). This protective effect was statistically significant after covariate adjustment, with the estimated hs-cTnT elevation rate among PART patients being 0.10 (95% CI: 0.02–0.26) compared with 0.35 (95% CI: 0.23–0.48) among other patients (all odds ratio *p*-values < 0.05).

Though the Bayesian adaptive sequential design requires substantial statistical computation, the clinical implementation of the PART framework is straightforward and seamlessly integrates into existing radiation oncology workflows. There are two models implemented in this study, one is the Bayesian NTCP model for personalized patient treatment, and the other is the BOP2 model for trial safety monitoring. The NTCP model generates specific MHD constraints (Table 2) for each risk group according to the risk profile. The personalized dose constraints can be directly used as planning directive to guide radiation treatment planning to reduce the MHD. Moreover, the MHD constraints are already familiar with radiation oncologist, dosimetrist and medical physicist, and can be easily implemented in clinical practice without additional training. The BOP2 model is used to determine the number of patients enrolled in each cohort and the go/no-go criteria for the trial (Table S1). The BOP2 monitoring enhances trial efficiency and ethical conduct by enabling early termination of ineffective interventions. The highlight of this design is that we will adapt the model when new evidence is available at each interim analysis instead of using a static model throughout the trial, providing a novel, data-driven approach to personalization. Such an adaptive design would enhance the predictive accuracy of the model, making it more robust and clinically applicable. As the trial progresses, we will further refine the model within the Bayesian framework, incorporating additional covariates (e.g., radiation modality, cardiac substructure doses) and stricter PART planning goals to optimize outcomes.

This trial has several limitations. First, this is a single-arm, single-institution study primarily designed as a proof of concept to demonstrate the feasibility of developing the PART model while simultaneously implementing it in patients through an adaptive trial. With more data and a refined PART model obtained from this trial, a future randomized trial would be of interest to formally establish the effectiveness of PART in a larger patient cohort. Second, most of the CAEs are late-onset, requiring a long observation window, which may delay timely futility monitoring. To address this, our trial incorporates the hs-cTnT level as a short-term surrogate endpoint in Bayesian monitoring alongside the 12-month grade ≥ 2 CAE. More advanced Bayesian monitoring designs^17,22^ that utilize time-to-CAE analysis to enhance monitoring efficiency could be implemented in future trials. Lastly, CAE is only one type of treatment-induced AEs in NSCLC patients. Thoracic chemoradiation exposes multiple organs at risk (e.g., lung, esophagus, and lymphoid tissue), all contributing to overall toxicity. We selected CAEs as the initial target AE to develop a prototype for the personalized therapy framework. In future work, we plan to extend the model by incorporating AEs from other organs at risk, given their potential interactions. Although our current intervention focuses on adapting the radiation dose to the heart rather than to the lung (i.e., the primary tumor), we do not anticipate PART to directly influence tumor-related mortality. However, by reducing cardiac risk, PART may lower cardiac event–related deaths and thereby indirectly contribute to a survival benefit. Taken together, these considerations highlight the importance of a more comprehensive risk–benefit assessment to inform clinical decision-making.

## CONCLUSION

Clinical implementation of PART model to guide treatment decision within a prospective trial is feasible. The recommended mean heart dose constraints generated by the first version of PART appear reasonable and clinically relevant. PART was associated with lower incidence of hs-cTnT elevation.

## Data Availability

Research data will be shared upon reasonable request to the corresponding author

## REFERENCES

1. Hardy D, Liu CC, Cormier JN, et al: Cardiac toxicity in association with chemotherapy and radiation therapy in a large cohort of older patients with non-small-cell lung cancer. Ann Oncol 21:1825–1833, 2010

2. Atkins KM, Rawal B, Chaunzwa TL, et al: Cardiac Radiation Dose, Cardiac Disease, and Mortality in Patients With Lung Cancer. J Am Coll Cardiol 73:2976–2987, 2019

3. Banfill K, Giuliani M, Aznar M, et al: Cardiac Toxicity of Thoracic Radiotherapy: Existing Evidence and Future Directions. J Thorac Oncol 16:216–227, 2021

4. Dess RT, Sun Y, Matuszak MM, et al: Cardiac Events After Radiation Therapy: Combined Analysis of Prospective Multicenter Trials for Locally Advanced Non-Small-Cell Lung Cancer. J Clin Oncol 35:1395–1402, 2017

5. Wang K, Eblan MJ, Deal AM, et al: Cardiac Toxicity After Radiotherapy for Stage III Non-Small-Cell Lung Cancer: Pooled Analysis of Dose-Escalation Trials Delivering 70 to 90 Gy. J Clin Oncol 35:1387–1394, 2017

6. Yegya-Raman N, Wang K, Kim S, et al: Dosimetric Predictors of Symptomatic Cardiac Events After Conventional-Dose Chemoradiation Therapy for Inoperable NSCLC. J Thorac Oncol 13:1508–1518, 2018

7. xxxxxxx

8. xxxxxxx

9. Karrison TG, Huo D, Chappell R: A group sequential, response-adaptive design for randomized clinical trials. Controlled Clinical Trials 24:506–522, 2003

10. Chow S-C, Song F: Adaptive clinical trial design. Quantitative Methods for HIV/AIDS Research:41–62, 2017

11. Apple FS, Jaffe AS, Collinson P, et al: IFCC educational materials on selected analytical and clinical applications of high sensitivity cardiac troponin assays. Clinical biochemistry 48:201–203, 2015

12. Cao L, Zhu W, Wagar EA, et al: Biomarkers for monitoring chemotherapy-induced cardiotoxicity. Critical reviews in clinical laboratory sciences 54:87–101, 2017

13. Lee JJ, Chu CT: Bayesian clinical trials in action. Stat Med 31:2955–72, 2012

14. Lin R, Lee JJ: Novel Bayesian Adaptive Designs and Their Applications in Cancer Clinical Trials, in Bekker A, Chen D-G, Ferreira JT (eds): Computational and Methodological Statistics and Biostatistics: Contemporary Essays in Advancement. Cham, Springer International Publishing, 2020, pp 395–426

15. Gelman A: Bayesian data analysis (ed Third edition.). Boca Raton, CRC Press, 2014

16. Zhou H, Lee JJ, Yuan Y: BOP2: Bayesian optimal design for phase II clinical trials with simple and complex endpoints. Stat Med 36:3302–3314, 2017

17. Lin R, Coleman RL, Yuan Y: TOP: Time-to-Event Bayesian Optimal Phase II Trial Design for Cancer Immunotherapy. J Natl Cancer Inst 112:38–45, 2020

18. Thall PF, Simon RM, Estey EH: Bayesian Sequential Monitoring Designs for Single-Arm Clinical-Trials with Multiple Outcomes. Statistics in Medicine 14:357–379, 1995

19. Siegel RL, Miller KD, Wagle NS, et al: Cancer statistics, 2023. Ca Cancer J Clin 73:17–48, 2023

20. Almatrafi A, Thomas O, Callister M, et al: The prevalence of comorbidity in the lung cancer screening population: A systematic review and meta-analysis. Journal of Medical Screening 30:3–13, 2023

21. Leduc C, Antoni D, Charloux A, et al: Comorbidities in the management of patients with lung cancer. European Respiratory Journal 49, 2017

22. Zhou H, Chen C, Sun L, et al: Bayesian optimal phase II clinical trial design with time-to-event endpoint. Pharm Stat 19:776–786, 2020

