## Supplementary-1 for "Bayesian Learning to Reduce Cardiac Risk for Locally Advanced NSCLC Patients Based on Personalized Radiotherapy Prescription"

**Supplementary material**

**S1. Derivation of the personalized radiation plan based on the personalized NTCP model**

A classification tree was developed for risk stratification based on tumor location and age, using input variables including tumor location, treatment modality, age, gender, race, Karnofsky performance score, smoking status, pre-existing heart disease (PHD), and baseline high-sensitivity cardiac troponin T (hs-cTnT) levels (BTnT). The Gini impurity index was used as the splitting criterion. Variable importance is shown in Figure S1. The first split was based on tumor location, with Group 1 consisting of patients with tumors in the right lung. For patients with tumors in the left lung or mediastinum, a second split was determined by age: those under 64 years were classified as Group 2, and those aged 64 years or older as Group 3. The classification tree achieved an accuracy of 0.74. Bootstrapping validation was performed to assess bias in model performance due to overfitting. We repeated the model development on 1,000 bootstrap samples and tested the model performance on the bootstrap sample and the original sample, yielding an optimism-corrected accuracy of 0.71.

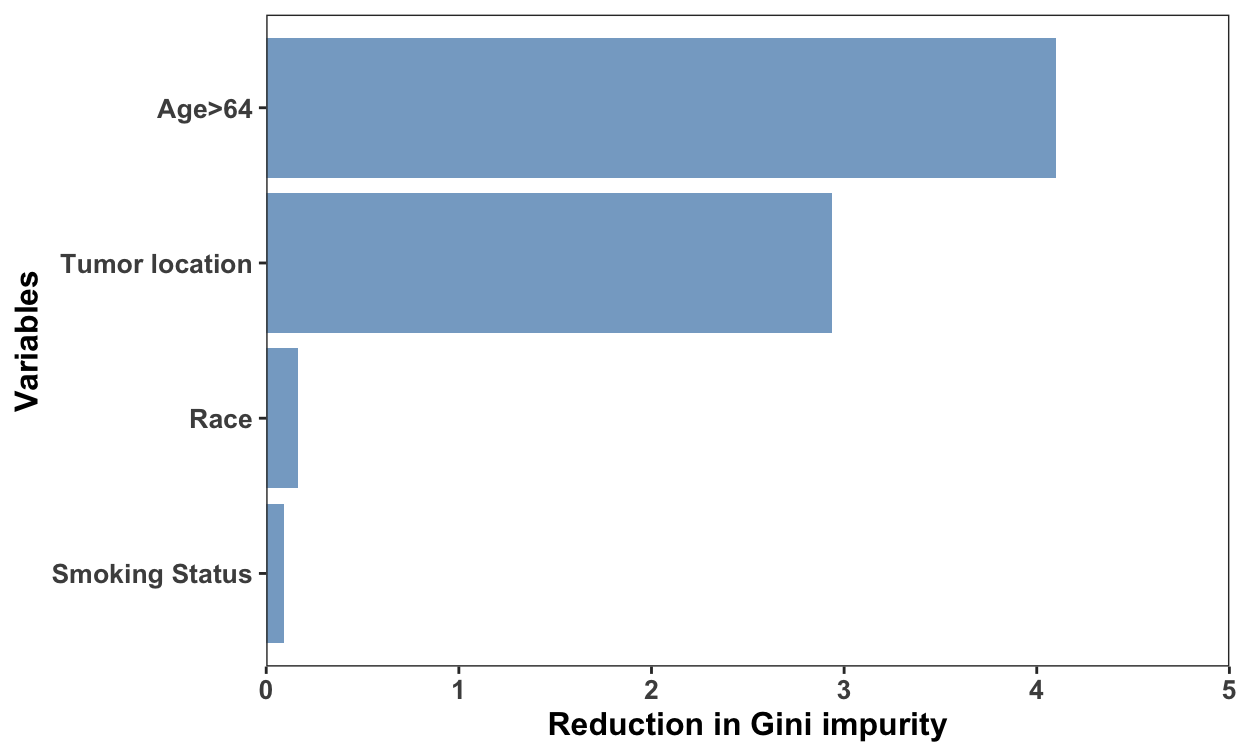

**Figure S1. Variable importance of the classification tree**

By using observed data and the chosen prior distribution for the unknown parameters, Bayesian posterior inference can be computed by using the well-known Bayes' theorem: posterior ∝ likelihood × prior. Many widely used Markov Chain Monte Carlo methods ^22^, such as the Gibbs sampler, can readily produce posterior samples of the unknown parameters. Assuming a total of $n$ patients have been treated and a total of $G$ risk groups are predefined, let $y_{i}=1$ indicate that the $i$th patient has experienced elevated hs-cTnT, and let $y_{i}=0$ indicate the absence of elevated hs-cTnT, $i=1,\ldots,n$. Let $p_{i}=\Pr\left( elevated hs-cTnT for patient i \right),$ then our full hierarchical logistic regression model is given by:

$$y_{i}\sim Bernoulli\left( p_{i} \right),$$

$$logit\left( p_{i} \right)=\alpha_{g_{i}}+\beta_{g_{i}}\log\left( MHD_{i} \right)+\gamma_{1}PHD_{i}+\gamma_{2}BTnT_{i},$$

$\alpha_{g}\sim N(\mu_{\alpha}, \sigma_{a})$, $\beta_{g}\sim N\left( \mu_{\beta}, \sigma_{\beta} \right), g=1,\ldots,G,$

where the subscript $i$ indicates the $i$th paitent. We take non-informative prior distributions for the parameters of the hierarchical logistic regression model as follows:

$$\mu_{\alpha}\sim N\left( 0, 100 \right),\mu_{\beta}\sim N\left( 0, 100 \right),$$

$$\sigma_{\alpha}\sim Unif\left( 0, 10 \right), \sigma_{\beta}\sim Unif\left( 0, 10 \right),$$

$$\gamma_{1}\sim N\left( 0, 100 \right), \gamma_{2}\sim N\left( 0, 100 \right).$$

Consequently, we can derive the posterior distribution of Pr(elevated hs-cTnT), given predetermined values for mean heart dose (MHD), PHD, BTnT, and the group indicator $g$. The predictive accuracy of the NTCP model had been evaluated using the area under receiver operating characteristic curve (AUC), and the median AUC is 0.725 (95% credible interval: 0.693-0.740) (Figure S2). In addition, the overfitting of the model has been assessed using Pareto-smoothed importance sampling leave-one-out cross-validation. All estimates for the shape parameter k of the generalized Pareto distribution were smaller than 0.7 and the effective number of parameters was smaller than the number of cases (7<160), indicating the model is well-specified with low risk of overfitting. Furthermore, we assessed alternative non-informative priors by varying the prior means and variances. The sensitivity analyses demonstrated that our model is robust to these choices, as the historical data dominate the Bayesian model fitting.

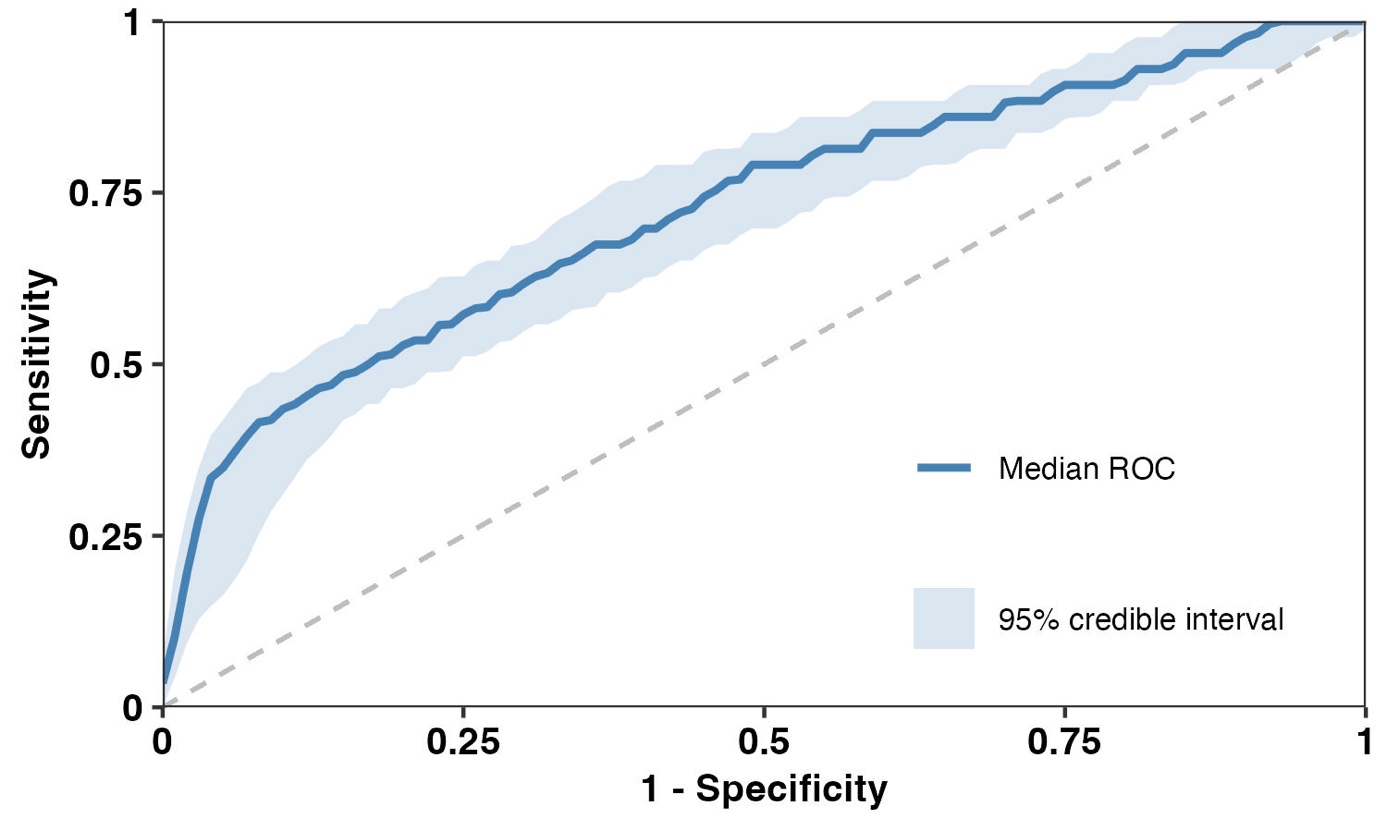

**Figure S2 Receiver operating characteristic curve of the Bayesian hierarchical logistic regression model**

In the first PART development, we first derived the posterior distribution of Pr(elevated hs-cTnT), given predetermined values for MHD, PHD, BTnT, and the group indicator. Let NTCPs represent the Pr(elevated hs-cTnT) given the MHD derived from the standard radiation plan; we then used the posterior probability Pr(NTCPs > X | Data) (denoted as PX) to assess whether the standard radiation plan is sufficiently safe for a given enrolled patient. Here, X represents the targeted probability of elevated hs-cTnT that we aim to achieve, and X can vary by group, indicating different targets for different groups. For example, we may consider setting a small X for low-risk groups and a larger X for high-risk groups. We carefully selected the value of X based on our historical trial data and cohort 1 data and through extensive discussions among investigators. In our initial PART development, X was chosen as 15%, 15%, and 45% for the three predefined risk subgroups (Table 3).

In fact, the quantity PX characterizes the posterior likelihood, based on the currently observed data, that the cardiac risk induced from the standard radiation plan is greater than the pre-specified target X. Compared with other commonly used statistics, e.g., the model point estimate of NTCPs using the maximum likelihood approach, PX not only quantifies the point estimate of NTCPs but also reflects the data-dependent credibility level (i.e., how confident we are in claiming that the standard radiation plan is overly toxic). If PX is sufficiently large, say greater than or equal to a probability threshold C (i.e., PX $\geq$ C), then the observed data suggest that the standard treatment plan is unacceptable because it yields a risk probability larger than X. On the other hand, if PX is small, say smaller than C (i.e., PX < C), then the standard treatment plan is deemed acceptable because the data suggest that it may yield a risk rate controlled below X. Therefore, the standard radiation plan can be retained for that specific patient. In cases where the PART strategy is deemed necessary, we once again refer to the posterior distribution of Pr(elevated hs-cTnT). We identify the highest dose that leads to a posterior probability of an elevated hs-cTnT rate exceeding X being smaller than C. The recommended MHD corresponding to this highest dose represents the recommended value for treating the patient. In clinical practice, when the planned MHD exceeded the constraints, dosimetrists were asked to re-optimize the plans to reduce the dose to the heart as much as possible without sacrificing the target coverage. Normally, the dose reduction was achieved by increasing the priority and decreasing the dose level of the generalized effective uniform dose optimization objective in the treatment planning system. This process is repeated until either the model’s recommendation is met or further reduction is not feasible without compromising tumor coverage.

In our trial, we optimized the cutoff probability C by minimizing the average loss from the point that achieves the best specificity and sensitivity in the subgroup-specific receiver operating characteristic curve. More specifically, let D denote the actual MHD received for the patient, and D_limit_(C) denote the highest MHD recommended by the above Bayesian model with PX smaller than C. The sensitivity refers to the ability that we can identify the patients whose MHD ($D)$exceeded the recommended dose limit D_limit_(C) (i.e., D>D_limit_(C)) and experienced elevation of hs-cTnT (Y=1).

$$Sensitivity(C)=\frac{N(D>D_{limit}(C),Y=1)}{N\left( D>D_{limit}(C),Y=1 \right)+N(D<D_{limit}(C), Y=1)}$$

The specificity is a measure of how well we can identify the patients whose MHD were below D_limit_(C) (i.e., D<D_limit_(C))and did not experience the event (Y=0).

$$Specificity(C)=\frac{N(D<D_{limit}(C),Y=0)}{N\left( D>D_{limit}(C),Y=0 \right)+N(D<D_{limit}(C), Y=0)}$$

The loss function is formulated as:

$$C_{optimal}=argmin\sqrt{{(1-Sensitivity\left( C \right))}^{2}+{(1-Specificity(C))}^{2}}$$

In other words, the optimal cutoff point in the ROC curve was selected based on the criterion that minimizes the Euclidean distance between sensitivity and 1-specificity and the optimal point (1,0).

**S2. Details of the BOP2 design**

Let $y_{1}$ denote the binary indicator of elevated hs-cTnT and $y_{2}$ denote the binary indicator of a grade $\geq2$ CAE. The joint outcome of $y_{1}$ and $y_{2}$ can be represented by a four-level multinomial variable $w$, where $w=1$ corresponds to $\left( y_{1},y_{2} \right)=(1,0)$, $w=2$ corresponds to $\left( y_{1},y_{2} \right)=(0,1)$, $w=3$ corresponds to $\left( y_{1},y_{2} \right)=(1,1)$, and $w=4$ corresponds to $\left( y_{1},y_{2} \right)=(0,0)$. Let $\theta_{k}=\Pr(w=k)$denote the probability of observing the $k$th level of $w$, $k=1,2,3,4$. As a result, the marginal probabilities can be represented as $p_{1}=\theta_{1}+\theta_{3}$ and $p_{2}=\theta_{2}+\theta_{3}$. Under the Bayesian framework, a Dirichlet prior $Dirichlet(\alpha_{1},\alpha_{2},\alpha_{3},\alpha_{4})$ is assigned to the joint probability vector $(\theta_{1},\theta_{2},\theta_{3},\theta_{3})$. In summary, based on data $D_{n}$ from $n$ patients, the Dirichlet-Multinomial model is expressed as

$$w_{i}\sim Multinomial\left( \theta_{1},\theta_{2},\theta_{3},\theta_{4} \right), i=1,\ldots,n,$$

$$\left( \theta_{1},\theta_{2},\theta_{3},\theta_{4} \right)\sim Dirichlet\left( \alpha_{1},\alpha_{2},\alpha_{3},\alpha_{4} \right).$$

Due to conjugacy, the posterior distribution of $\left( \theta_{1},\theta_{2},\theta_{3},\theta_{4} \right)\mid D_{n}$ remains a Dirichlet distribution, and is given by $\left( \theta_{1},\theta_{2},\theta_{3},\theta_{4} \right)\mid D_{n}\sim Dirichlet\left( \alpha_{1}+\sum_{i=1}^{n} 1\left( w_{i}=1 \right),\alpha_{2}+\sum_{i=1}^{n} 1\left( w_{i}=2 \right),\alpha_{3}+\sum_{i=1}^{n} 1\left( w_{i}=3 \right),\alpha_{4}+\sum_{i=1}^{n} 1\left( w_{i}=4 \right) \right),$ where $1\left( \cdot\right)$ is the indicator function. Due to the properties of the Dirichlet distribution, the posterior distributions of $p_{1}\mid D_{n}$ and $p_{2}\mid D_{n}$ follow Beta distributions: $p_{1}\mid D_{n}\sim Beta(\alpha_{1}+\alpha_{3}+\sum_{i=1}^{n} 1\left( w_{i}=1 or w_{i}=3 \right), \alpha_{2}+\alpha_{4}+\sum_{i=1}^{n} 1\left( w_{i}=2 or w_{i}=4 \right))$ and $p_{2}\mid D_{n}\sim Beta(\alpha_{2}+\alpha_{3}+\sum_{i=1}^{n} 1\left( w_{i}=2 or w_{i}=3 \right), \alpha_{1}+\alpha_{4}+\sum_{i=1}^{n} 1\left( w_{i}=1 or w_{i}=4 \right))$.

Let $p_{10}$ and $p_{20}$ denote the historical control rates. The null hypothesis $H_{0}:\left( \theta_{1},\theta_{2},\theta_{3},\theta_{4} \right)=\left( \theta_{10},\theta_{20},\theta_{30},\theta_{40} \right)$ assumes independence between $y_{1}$ and $y_{2}$, which implies $\theta_{10}=p_{10}(1-p_{20})$, $\theta_{20}=p_{20}(1-p_{10})$, $\theta_{30}=p_{10}p_{20},$ $\theta_{40}=(1-p_{10})(1-p_{20})$. By incorporating this formulation, the resulting BOP2 design becomes more robust in controlling the type I error rate when $y_{1}$ and $y_{2}$ are positively correlated. By default, the BOP2 design adopts a pessimistic approach in selecting the Dirichlet prior, where $\left( \alpha_{1},\alpha_{2},\alpha_{3},\alpha_{4} \right)$ are set as $\left( \theta_{10},\theta_{20},\theta_{30},\theta_{40} \right)$. Thus, the effective sample size of the prior is $\alpha_{1}+\alpha_{2}+\alpha_{3}+\alpha_{4}=1$. In our study, we take $p_{10}=0.25$, $p_{20}=0.15$, $\theta_{10}=0.2125$, $\theta_{20}=0.1125$, $\theta_{30}=0.0375, \theta_{40}=0.6375$.

Based on the Beta posteriors of $p_{1}\mid D_{n}$ and $p_{2}\mid D_{n}$, we will stop enrolling patients and declare PART not effective if there is a small posterior probability of achieving a rate of hs-cTnT elevations smaller than 25% and a rate of CAEs smaller than 15%, that is:

$$\Pr\left( p_{1}<0.25 | Data \right)<\lambda\left( \frac{n}{N} \right)^{\alpha}, \mathrm{and} \Pr\left( p_{2}<0.15 | Data \right)<\lambda\left( \frac{n}{N} \right)^{\alpha}.$$

Here, $N=200$ is the maximum sample size, $n$ is the interim sample size. The design parameters of BOP2, i.e., $\lambda$ and $\alpha$, are then optimized to maximize the probability of correctly concluding that an effective PART plan is acceptable under the alternative hypothesis $H_{1}$ (i.e., the study power), while controlling that the probability of incorrectly claiming an ineffective PART plan as acceptable is less than 5% under the null hypothesis $H_{0}$ (i.e., the type I error rate). In this study, we would consider PART to be effective if $p_{1}=0.15$ and $p_{2}=0.10$ under $H_{1}:\left( \theta_{1},\theta_{2},\theta_{3},\theta_{4} \right)=(0.135, 0.085, 0.015, 0.765)$, i.e., a decrease of 0.1 in the probability of elevated hs-cTnT or a decrease of 0.05 in the probability of CAE.

The optimized design can be obtained using the web-based shiny application at <https://trialdesign.org/one-page-shell.html#BOP2> by selecting the 'Multiple Efficacy' option and setting efficacy as 1 – toxicity. The optimized values of $\lambda$ and $\alpha$ are $\lambda=0.98$ and $\alpha=1$, yielding the study power of 0.969 and the type I error rate of 0.043.

**S3. Comparison of hs-cTnT outcomes between cohort 1 and cohort 2**

We report a comparison of hs-cTnT outcomes between cohort 1 and cohort 2 using three methods:

(1) Raw comparison: We fitted a logistic regression model with only an intercept and the cohort indicator. The odds ratio and its 95% confidence interval were derived.
(2) Inverse propensity weighting (IPW)-adjusted comparison: We estimated the propensity score using a logistic regression model that treated cohort assignment as the “treatment” and included covariates such as baseline hs-cTnT level, age, sex, tumor location, pre-existing heart disease, and treatment modality. The standardized mean difference (SMD) was used to assess balance between the two cohorts after IPW, and the results (Table S3) showed that the cohorts were well balanced. After IPW, the average treatment effect was estimated by fitting a weighted logistic regression, from which the odds ratio and its 95% confidence interval were derived.
(3) Regression-adjusted comparison: We fitted a logistic regression model including the cohort indicator along with baseline hs-cTnT level, age, sex, tumor location, pre-existing heart disease, and treatment modality. From this model, we obtained the marginal odds ratio and its 95% confidence interval.

As nine patients had missing hs-cTnT elevation data, we implemented multiple imputation using the mice package in R to generate 100 imputed datasets for the three comparison methods. Specifically, we applied predictive mean matching with patient baseline and modality variables as predictors. Rubin’s rules were then used to combine results and calculate the variance of the estimated statistics across the imputed datasets.

Across the three analytic approaches, Table 3 indicates that patients in cohort 2 consistently showed a lower odds of hs-cTnT elevation compared with cohort 1. The raw comparison yielded an odds ratio of 0.47 (95% CI: 0.19–1.16, p = 0.101). After adjustment using inverse propensity weighting, the estimated odds ratio was 0.41 (95% CI: 0.16–1.04, p = 0.060), suggesting a trend toward reduced risk in cohort 2 with improved covariate balance. The regression-adjusted comparison produced a similar odds ratio of 0.46 (95% CI: 0.17–1.29, p = 0.139). Although none of the comparisons reached conventional statistical significance, the results were directionally consistent across methods, indicating a potential reduction in hs-cTnT elevation in cohort 2 relative to cohort 1.

**S4. Comparison of hs-cTnT outcomes between patients meeting versus exceeding PART dose constraints in cohort 2**

We also compared hs-cTnT outcomes between patients who met the PART dose constraints (“PART patients”) and those who exceeded the constraints (“Non-PART patients”) within cohort 2, using the same three methods described in Section S3. Standardized mean differences (SMDs) of the selected variables before and after IPW adjustment are reported in Table S3, and Table 3 summarizes the odds ratio estimates.

The results in Table 3 consistently suggested a lower incidence of hs-cTnT elevation among PART patients compared with Non-PART patients. The raw comparison yielded an odds ratio of 0.28 (95% CI: 0.06–1.18, p = 0.083). After IPW adjustment, the odds ratio was further reduced to 0.10 (95% CI: 0.02–0.62, p = 0.013), demonstrating significant covariate balance and a strong protective effect of meeting PART constraints. Similarly, the regression-adjusted comparison produced an odds ratio of 0.11 (95% CI: 0.01–0.84, p = 0.033). Taken together, these results indicate that patients whose treatment satisfied the PART dose constraints had a substantially lower risk of hs-cTnT elevation relative to those whose treatment exceeded the constraints in cohort 2.

**S5. Comparison of hs-cTnT outcomes between patients meeting PART dose constraints in cohort 2 and other patients**

We also compared hs-cTnT outcomes between patients who met the PART dose constraints (“PART patients”) in cohort 2 and a combined group of patients who exceeded the PART dose constraints in cohort 2 together with all patients in cohort 1 (“Other patients”). The same three methods described in Section S3 were applied. Standardized mean differences (SMDs) of the selected variables before and after IPW adjustment are reported in Table S3, and Table 3 summarizes the odds ratio estimates.

The results in Table 3 consistently showed a lower risk of hs-cTnT elevation among PART patients compared with other patients. The raw comparison yielded an odds ratio of 0.29 (95% CI: 0.09–0.86, p = 0.026). The IPW-adjusted comparison further supported this finding, with an odds ratio of 0.25 (95% CI: 0.08–0.82, p = 0.022). Similarly, the regression-adjusted comparison produced an odds ratio of 0.16 (95% CI: 0.04–0.65, p = 0.010). Collectively, these results suggest that patients whose treatment satisfied the PART dose constraints experienced a substantially lower incidence of hs-cTnT elevation compared with those who did not.

**Table S1 Optimized stopping boundaries used in monitoring the effectiveness of the developed personalized adaptive radiation treatment (PART)**

| No. of patients treated | Stop if both conditions are satisfied | |
| --- | --- | --- |
|  | **No. of elevated hs-cTnT is ≥** | No. of cardiotoxicities is ≥ |
| 50 | 15 | 10 |
| 100 | 26 | 16 |
| 150 | 35 | 21 |
| 200 | 38 | 21 |

**Table S2** **Summary statistics for evaluable patients in terms of hs-cTnT elevation of the first two cohorts in this Bayesian adaptive cohort registration trial as of July 31, 2025.**

|  | **Cohort 1** | **Cohort 2** | **P Value** |
| --- | --- | --- | --- |
| **No. of evaluable patients** | 47 | 44 |  |
| **No. of events** | 15 (32%) | 9 (20%) | 0.316 |
| **Age, y, mean ± SD** | 65.23 ± 9.33 | 68.68 ± 9.42 | 0.073 |
| Age ≥64 | 27 (57%) | 32 (73%) | 0.192 |
| <64 | 20 (43%) | 12 (27%) |  |
| **Sex** |  |  | 0.565 |
| Male | 22 (47%) | 17 (39%) |  |
| Female | 25 (53%) | 27 (61%) |  |
| **Tumor location** |  |  | 0.616 |
| Right | 27 (57%) | 22 (50%) |  |
| Left/mediastinal | 20 (43%) | 22 (50%) |  |
| **Risk groups** |  |  | 0.530 |
| Group 1: Tumor location = right | 27 (57%) | 22 (50%) |  |
| Group 2: Tumor location = left/mediastinal, age <64 | 8 (17%) | 6 (14%) |  |
| Group 3: Tumor location = left/mediastinal, age ≥64 | 12 (26%) | 16 (36%) |  |
| **Pre-existing heart disease** |  |  | 1.000 |
| Yes | 17 (36%) | 16 (36%) |  |
| No | 30 (64%) | 28 (64%) |  |
| **Baseline hs-cTnT level, mean ± SD** | 9.86 ± 8.37 | 16.78 ± 32.7 | 0.133 |
| >10 ng/L | 16 (34%) | 22 (50%) | 0.184 |
| ≤10 ng/L | 31 (66%) | 22 (50%) |  |
| **Treated mean heart dose, Gy, mean ± SD** | 7.84 ± 6.26 | 5.74 ± 5.7 | 0.079 |

Abbreviations: SD=standard deviation, hs-cTnT=high-sensitivity cardiac troponin T assay

**Table S3 Standardized Mean Difference (SMD) before and after IPW**

| **Measure** | **Type** | **Unadjusted SMD** | **Adjusted SMD** |
| --- | --- | --- | --- |
| (1) **Comparing Patients in Cohort 1 and Cohort 2** | | | |
| Propensity Score | Distance | 0.5390 | 0.0676 |
| Baseline hs-cTnT | Continuous | 0.2711 | 0.1048 |
| Pre-existing heart disease | Binary | 0.0200 | 0.0253 |
| Sex | Binary | -0.0800 | -0.0033 |
| Tumor location | Binary | -0.1000 | -0.0150 |
| Age | Continuous | 0.3481 | 0.0289 |
| Modality | Binary | 0.1000 | 0.0107 |
| (2) **Comparing PART and Non-PART Patients Within Cohort 2** | | | |
| Propensity Score | Distance | 1.8292 | 0.2159 |
| Baseline hs-cTnT | Continuous | -0.5582 | -0.2001 |
| Pre-existing heart disease | Binary | -0.3603 | 0.0445 |
| Sex | Binary | -0.2684 | -0.0185 |
| Tumor location | Binary | -0.2757 | -0.2022 |
| Age | Continuous | -0.5009 | -0.2161 |
| Modality | Binary | 0.3676 | 0.0060 |
| (3) **Comparing PART in Cohort 2 and Other Patients in Cohorts 1&2** | | | |
| Propensity Score | Distance | 0.8415 | 0.0951 |
| Baseline hs-cTnT | Continuous | -0.2680 | -0.1342 |
| Pre-existing heart disease | Binary | -0.1595 | -0.0350 |
| Sex | Binary | -0.1907 | -0.0839 |
| Tumor location | Binary | -0.2094 | -0.0373 |
| Age | Continuous | 0.0190 | 0.0392 |
| Modality | Binary | 0.2540 | 0.0180 |

Abbreviations: hs-cTnT=high-sensitivity cardiac troponin T assay, CI=confidence interval, IPW=Inverse probability weighting; PART = personalized adaptive radiation treatment
