## Supplementary-2 for "Bayesian Learning to Reduce Cardiac Risk for Locally Advanced NSCLC Patients Based on Personalized Radiotherapy Prescription"

**Longitudinal Assessment of Cardiovascular Injury and Cardiac Fitness in LA-NSCLC Patients Receiving Model Based Personalized Chemoradiation – an Adaptive Cohort Registration Study (ClinicalTrials.gov Identifier: NCT05010109)**

**PI**: Zhongxing Liao, MD, Department of Radiation Oncology

1.0 **Objectives:**
    1.1 **Overall objective:** To longitudinally assess cardiac injury (serum biomarkers, and grade ≥2 cardiac events), and overall cardiac fitness (6-minute-walk test) in LA-NSCLC patients receiving chemoradiation. The radiation modality and technique will be selected by personalized multivariable model prediction as the optimal with the lowest possible cardiac toxicity based on the individual risk while maintaining standard dosimetric constraints for the tumor and other normal tissues. The long terms goal is to improve outcome of lung cancer patient by reducing the cardiac toxicity.
    1.2 **Primary endpoints:** 1). Increase from baseline in level of hs-TnT >=5ng/L at the end of CRT; or 2) Grade ≥2 cardiovascular events defined by CTCAE v5.0 (Appendix 1) within 12-month of completion of CRT (https://ctep.cancer.gov/protocoldevelopment/electronic_applications/docs/ctcae_v5_quick_reference_5x7.pdf).
    1.3 **Secondary endpoints:**
        1.3.1 Assess the trajectory of overall cardiac fitness and functional capacity and impact of radiation exposure using 6MWT and correlation with primary endpoints
        1.3.2 Assessing trajectory of patient reported outcomes using EQ- 5D-5L and MDASI-LC and correlation with primary endpoints
        1.3.3 Assess the association between spatial radiation dose distribution and 3D cardiac injury on SPECT/CT, cardiac PET- images, and serial EKGs.
        1.3.4 Determine the trajectory and association of serum cardiac biomarker levels with radiation exposure
        1.3.5 Long term cardiovascular events
        1.3.6 OS and PFS

**2.0 Rationale**

Currently, there are 14 million cancer survivors in the United States and this number is anticipated to exceed 20 million by 2026, resulting from innovation and improvements in systemic (e.g., immunotherapy) and local therapies (e.g., proton therapy).^1^ However, this improvement achievement is tempered by significant treatment-related toxicities. Lung cancer is the most common cancer worldwide that accounts for approximately 25% of all cancer-related deaths.^1^ For patients with surgically unresectable locally advanced NSCLC, the standard of care is chemotherapy and/or immunotherapy combined with radiotherapy. Even though lung cancer survival has improved over time,^2^ treatment-related toxicities can have both short- and long-term consequences on the morbidity and mortality of NSCLC patients.

Cardiac toxicity has long-been observed as an adverse effect to traditional photon radiotherapy in NSCLC patients.^3^ The reported incidence of symptomatic cardiac events is as high as 28.6% in NSCLC patients and occurs as early as 3 months after chemoradiation,^4^ differing from those reported in breast and lymphoma patients, wherein most of the cardiac toxicity occurs years after treatment.^5^ Cardiac events and overall survival (OS) are associated with mean heart dose (MHD) and subvolume doses^6,7^ and doses to specific anatomical cardiac substructures.^4^ Advancements in radiotherapy technologies such as volumetric modulated arc therapy (VMAT) and passively scattered proton therapy (PSPT) have been used to reduce cardiac exposure to radiation. However, exposure and damage to the heart is inevitable due to the close proximity of lung tumors to heart. In our first randomized trial, PSPT improved MHD;^8^ however, reduced MHD did not translate into reduced cardiac toxicity. Intensity modulated proton therapy (IMPT) could shape proton beam to better protect the heart, but substructures of the heart may still receive high doses, which are strongly associate with survival.^9^ We observed that cardiac toxicity could occur even when MHD was low, and only a portion of patients developed cardiac events for a given MHD, suggesting there exist interpatient differences in susceptibility to cardiac toxicity. Furthermore, lung cancer patients often present with factors, such as preexistent cardiac disease, smoking-related comorbidities, diabetes, etc. How these factors influence cardiac toxicity has not been fully studied. Thus, it is imperative to characterize patient and clinical factors to identify those patients for whom no safe dose threshold exists before cancer treatment starts.

The dose distribution patterns of photon and proton distribution in patients are dramatically different. For a given mean dose, photons will have a large, low and intermediate dose ‘bath’, whereas protons, especially passively scattered protons, may have a larger, high-dose volume adjacent to the target even though the low dose bath is significantly smaller. However, for heart, high dose, even to a small volume (e.g., substructure), may cause more injury and be more clinically relevant. In a previous study, comparing cardiac substructure exposure from IMRT vs. proton beam therapy (PBT) in esophageal cancer patients, the authors found that PBT resulted in significantly lower radiation exposure to cardiac substructures than IMRT.^10^ In general, however, in the current practice, dose to substructures of the heart are not considered in radiation planning. Moreover, our knowledge of dosimetric tolerances of heart substructures is poor.

Radiation dose limit to a specific volume of an organ (dose-volume constraints) is commonly used to determine if a radiotherapy plan is safe for the patient. Dose-volume parameters are usually used in normal tissue complication probability (NTCP) models for predicting risk of normal tissue toxicity. The Lyman–Kutcher–Burman (LKB) model is the best-known NTCP model. We tested the LKB NTCP model to predict radiation-induced esophagitis in NSCLC patients receiving PSPT and found that this model could predict grade ≥ 2 radiation esophagitis.^11^ LKB NTCP model was also used to determine the incidence of acute radiation-induced dermatitis in thoracic cancer patients treated with IMRT or PSPT on a completed prospective randomized trial.^12^ However, there are three major shortcomings of current models (including the LKB model) when applied to predict cardiac toxicity: 1) all cardiac toxicity models in the literature are based on photon radiotherapy data; 2) the current models usually consider the heart as a uniform organ without differences in vulnerability of substructures or functional subunits; and 3) models are population-based without considering an individual’s risk factors. Thus, current models for designing radiotherapy plans may not be accurate in assessing safe radiation doses for the whole heart and its complex substructures. A more advanced model needs to be created to take into consideration patient-specific genetic-, cardiac-, systemic therapy-, and radiotherapy-related factors.

Pre-existing cardiovascular risk factors (e.g., coronary artery calcium [CAC] score) have been used to assess long-term outcomes, such as all-cause mortality, in patients upon lung cancer screening. CAC scores are typically non-invasive measurements of coronary atherosclerosis and, also indicators of the severity of cardiac adverse event.^13^ CAC can be detected using computed tomography (CT) and scoring algorithms are in place for both cardiac-gated and non-cardiac-gated studies, which have a high level of agreement.^14^ The cardiac co-Investigators (co-I), Drs.Anita Deswal and Efstratios Koutroumpakis, on this proposal has previously shown that CAC scores are strong predictors of cardiac events, specifically among women, despite a low baseline traditional risk factor burden.^15^ CAC progression is considered a prognostic tool for accelerated, long-term coronary atherosclerosis and subsequent cardiac events; however, whether CAC scores can predict short-term outcomes (2-3 years), in particular, in response to chemoradiation, is not known.

In addition to CAC scores, cardiac injury biomarkers, such as high-sensitivity cardiac Troponin T [hs-cTnT], Brain Natriuretic Peptide (BNP), and C-reactive protein (CRP) also predict a higher likelihood of myocardial ischemia. Several studies have demonstrated that hs-cTnT assays enable the early detection and prediction of chemo- or radiotherapy-induced cardiac toxicity.^16-20^ In fact, our previously funded study showed that hs-cTnT levels began increasing as early as week 2 during the chemoradiation therapy (CRT) regimen, and hs-cTnT elevation at the time of pre-treatment, during treatment, and post-treatment was predictive of cardiac events and 2-year OS in NSCLC patients. In cancer patients receiving radiotherapy, a meta-analysis suggested that BNP could be used as a biomarker of cardiac damage.^9^ Another marker of cardiac damage from inflammation and ischemia is CRP, which has been shown to effectively monitor trastuzumab-induced cardiac toxicity in early-stage breast cancer,^21^ and is associated with severity of thoracic radiotherapy-induced cardiomyopathy.^22^

Cardiac damage from CRT may also be predicted by other pre-existing cardiovascular risks, such as the 10-year and lifetime risks for atherosclerotic cardiovascular disease (ASCVD). ASCVD risk includes age, sex, race, total cholesterol, high-density lipoprotein (HDL) cholesterol, systolic blood pressure, blood pressure medication use, diabetes status, and smoking status. Not only may ASCVD help predict outcomes to chemoradiation, but also genetic differences between the NSCLC patients may be predictive of outcomes. Indeed our previous studies showed genetic single nucleotide polymorphisms (SNPs) in genes involved in DNA repair and inflammation were associated with chemoradiation-induced pneumonitis.^23,24^ In preclinical studies, differences in mitochondrial gene expression caused by inherited genetic variants contributed to differences in sensitivity to cardiac radiation.^25^

The damage to the heart after CRT can lead to left ventricular (LV) dysfunction. Although LV dysfunction is typically determined by cardiac ejection fraction (EF), EF does not reflect regional differences in LV function and myocardial deformation (i.e., strain). Myocardial strain is measured using speckle tracking echocardiography, and cardiac radiation doses administered to breast cancer patients have been shown to be associated with changes in myocardial strain.^26^ However, the study proposed in this application will be this first to use speckle tracking echocardiography to correlate myocardial strain with radiation doses to heart and its substructures in NSCLC patients, thus, filling a significant gap in knowledge.

The ability of NSCLC patients to physically function and exercise is of prognostic significance.^27^ Measurement of functional capacity in NSCLC patients is commonly determined by 6-minute walk test (6MWT) (American Thoracic Society guideline). A 6MWT is a valid cardiac fitness test of functional capacity in cancer patients,^28^ with a 10% decline being a clinically relevant change.^29^ 6MWT has been show to predict radiation-induced pulmonary toxicity in combination with forced expiratory volume in 1 second.^30^ However, less is known regarding the prognostic value of functional capacity to predict adverse cardiac toxicity following radiotherapy for lung cancer. In addition, whether altering cardiac dose and/or treatment modality (photon versus proton) positively impacts functional capacity in lung cancer patients is currently unknown. Moreover, patient reported outcomes (PRO)s are powerful tool to inform decision-makers about patient morbidity and could improve the quality of the care.

We propose to characterize patients who are at high risk for cardiac toxicity, develop a multivariable model to guide personalized dosimetric constraints to reduce heart injury, and longitudinally assess the cardiac injury and function and functional capacity of NSCLC patients after chemoradiation therapy. Specifically, the proposed research aims to develop accurate and clinically useful multivariable models for cardiac toxicity, integrating relevant clinical, biological, and dosimetry factors which will allow clinicians to prescribe radiation treatment customized to individual risk. Our overarching goal is to minimize cardiovascular injury while optimizing NSCLC outcomes after concurrent CRT using a multivariable model to select radiation therapy modality and technique based on individual’s risk of cardiac injury. Currently, there is no methodology to help physicians identify patients who would be at serious risk of developing cardiac events to guide treatment design. The proposed project will lead to significant advancement in the personalization of radiotherapy. Furthermore, because the design of radiation therapy follows a set of common principles regardless of disease site, this research could benefit hundreds of thousands of cancer patients since at least 50% of the 1.8 million estimated new cancer patients in the U.S. each year will need radiation as a component of curative multidisciplinary treatment.^31^

**3.0 Eligibility of Subjects** Inclusion criteria:

1. Patient with histologic diagnosis of non-small cell lung cancer, small cell lung cancer, or limited stage – small cell lung cancer (L-SCLC)
2. The recommended treatment is thoracic radiation therapy combined with concurrent systemic therapy (chemotherapy and/or immunotherapy) with or without neoadjuvant and/or adjuvant systemic therapy (chemotherapy, immunotherapy, targeted therapy)
3. >/= 18 years of age
4. KPS >/= 70
5. Willing and able to sign informed consents
6. Willing to perform 6minute walking test
7. Willing to preform required cardiac biomarker test for primary end point assessment.

Exclusion criteria:

1. Unable or unwilling to give written informed consent
2. Previous history of RT to the thorax overlapping with the current treatment field.
3. Pregnant or breast-feeding
4. Renal failure necessitating dialysis
5. Unwilling to perform protocol tests
6. Contraindication for any protocol tests

**4.0 Research Plan and Methods**

This is a prospective adaptive cohort registration study of patients who will receive RT that involves the delivery of dose to the heart. As a part of the study, patients will be evaluated by myocardial perfusion SPECT/CT with stress test, echocardiogram with strain, and 6 MWT. Blood and urine samples will be collected. These evaluations will not influence their cancer treatment. The procedures will involve no more than minimal risk (i.e. rare complications related to echocardiography, SPECT/CT, or venipuncture).

**4.1 Schedule of data collection Table 1:**

| Variables | Before RT | RT  wk1 | RT wk2-3 | RT  wk4-5 | RT wk6-7 | 6-8 wks after RT | 4-6 mo after RT | 12 mo after RT | Annually 24 mo after RT |
| --- | --- | --- | --- | --- | --- | --- | --- | --- | --- |
| Cardiac History | X |  |  |  |  |  | X | X | X |
| Peripheral leukocyte DNA for SNP | X |  |  |  |  |  |  |  |  |
| *Ca^++^ scores | X |  |  |  |  |  | X | X | X |
| Cardiac RT dose |  | X | X | X | X |  |  |  |  |
| Immunotherapy | X |  |  |  |  |  | X | X |  |
| PRO | X |  | X | X | X | X | X | X | X |
| **Serum/plasma (hsTnT, NT-proBNP, hs-CRP, CK-MB) | X |  | X | X | X | X | X | X | X |
| ***ASCVD score | X |  |  |  |  |  |  |  |  |
| 6-minute walk | X |  | X |  | X | X | X | X |  |
| EKG | X |  | X |  | X | X | X | X |  |
| #Echocardiogram strain | X |  |  |  |  | X |  | X |  |
| ##Stress SPECT/CT | X |  |  |  |  | X |  | X |  |
| ### Pulmonary Function Test | X |  |  |  |  | X | X | X | X |

*, **, *** If these tests have been performed as part of another trial or as part of SOC and results are available, these tests should not be repeated.

#,##, ###Optional cardiac imaging studies. If the Stress SPECT/CT and/or 2D Echo with strain have been performed as part of the standard of care, these cardiac images should not be repeated. Outside cardiac imaging results are acceptable as well. The baseline imaging could be done before the 5^th^ fraction of radiation therapy.

**4.2 Patient enrollment:**

This protocol will follow the SOP 04_Informed Consent Process. SOP 04 has been read by the research staff and investigators. Informed consent may be obtained using remote consent.Appropriate patients will be identified in the Radiation Oncology clinic and offered enrollment on protocol by the treating physician. Patients who enrolled to 2020-0995 are eligible for this protocol as well. All enrolled patients will provide written informed consent.

**4.3 Patient history and ASCVD score ^32^**

Patients will undergo routine preclinical evaluation including the liver, renal functional tests, Lipid panel, A1C, and detailed cardiac history as part of the standard of care for lung cancer pretreatment evaluation. If medically indicated, patient will be referred to cardiology for evaluation.

**4.4 Myocardial Perfusion SPECT/CT**

This is optional imaging test offered to patient who does not have clinical indication for this examination. We will utilize attenuation corrected single photon emission computed tomography (SPECT)/CT cardiac perfusion imaging as opposed to qualitative SPECT in the previous research ^33,34^. We will register the CT images of SPECT/CT with the simulation CT images in the radiation treatment plan. The SPECT/CT will be performed with either exercise or pharmacological stress unless patients cannot tolerate the stress test. Patients will undergo stress Cardiac SPECT/CT according to the current standard clinical workflow.

If medically indicated, we will also refer patients to have Cardiac PET/CT

**4.5 Echocardiography with strain measurements:**

Echocardiograms will be performed by experienced sonographers using a GE machine (Vivid 7; GE Healthcare, Milwaukee, WI, USA) with a 2.5 or 3.5 MHz phased array transducer. 2D and color Doppler images from the apical four-, three- and two-chamber views will be acquired, and all echocardiographic measurements will be performed offline in a blinded fashion. Quantitative LVEF will be measured using the Bi-plane Simpson’s Method. Diastolic parameters including early mitral inflow velocity (E), the late diastolic velocity (A), the deceleration time and, the E/A ratio, the systolic velocity of the medial mitral annulus (Smed), the early diastolic velocity of the medial mitral annulus (E’med), the late diastolic velocity of the medial mitral annulus (A’med), the systolic velocity of the lateral mitral annulus (Slat), the early diastolic velocity of the lateral mitral annulus (E’lat), and the late diastolic velocity of the lateral mitral annulus (A’lat) will be measured. Global longitudinal strain will be measured using 2D speckle-tracking echocardiography with 3 long axis images obtained at high frame rates (50-70fps). Analysis will be performed using the EchoPAC software (version BT11, GE Medical, Milwaukee, WI, USA).

This is an optional examination offered to patients enrolled to this trial. Per ASE guidelines, the serial Echo Cardiography with strain prior RT, just after RT, and annually after is considered as standard of care ^35,36,37^. Variables of interest include: ejection fraction, strain, strain rate, ventricular-arterial coupling, diastolic function, left atrial volume.

**4.6 Electrocardiogram:**

Electrocardiograms will be performed to assess for arrhythmias as standard of care.

**4.7 Coronary artery calcium scores:**

As an exploratory analysis, coronary artery calcium scores will be assigned to the simulation CT and to CT scans that are performed as a part of routine patient follow-up after RT. Scores from before and after RT will be compared to assess for coronary artery calcification after the completion of RT. As indicated medically, patients may have a dedicated nuclear myocardial perfusion SPECT/CT or cardiac FDG PET/CT scan.

**4.8 Laboratory procedures:**

Blood will be collected within 2 weeks prior to RT, 2-3 weeks after the initiation of RT, during the final week of RT, at 3, 6, 12, and 24 months after completion of RT. Whenever possible, blood will be drawn at the time of regularly scheduled laboratory evaluations, so it will not involve an additional procedure. For samples taken during the course of RT, blood will be drawn after that day’s fraction, whenever possible.

At each interval, the following blood samples will be collected: ·

Blood specimens:
2 (10mL) “red” top tube (No Additive)
2 (10 mL) green top tube

3 (9 mL) lavender stopper (K2 EDTA)

Patients enrolled on the LAB09-0307 protocol will have signed a separate consent for blood that is drawn for that study in the appropriate tubes and at the appropriate time points can be used for analyses in this study. It is not necessary to draw additional, identical tubes. Serum will be stored as frozen specimens in locked storage in the laboratory of a research team member.

All blood biomarkers will be analyzed in the clinical chemistry core laboratories. Blood samples will be collected into lithium heparin tubes and centrifuged. The plasma is removed and stored at -80°C until analysis. The hs-cTnT concentrations are measured with highly sensitive cTnT reagents on Roche cobas 8000 automated analyzer using electrochemiluminescence immunoassay (Roche Diagnostics, Indianapolis, IN, USA). NT-proBNP is measured also using electrochemiluminescence immunoassay on the same Roche analyzer. High sensitivity CRP (hsCRP) in plasma is measured using particle enhanced immunoturbidimetric assay on Roche cobas® e411 automated analyzer (Roche Diagnostics, Indianapolis, IN, USA).

Table 2 list the **frequently used biomarkers of cardiotoxicity (Table 2)**

| **Full name** | **Abbreviation** | **Clinical relevance when elevated** |
| --- | --- | --- |
| Brain-type Natriuretic Peptide | NT-proBNP | Hypernatremia, high blood volume or pressure |
| Cardiac Troponin T | hs-TnT | Myocardial injury or necrosis |
| Creatinine kinase isoform MB | CK-MB | Myocardial injury or necrosis |
| High sensitivity C-reactive protein | hsCRP | Inflammation |
| Growth differentiation factor | GDF-15 | Inflammation and oxidative stress |
| Myeloperoxidase | MPO | Oxidative stress |
| Placental growth factor | PIGF | Increasing angiogenesis |
| Soluble fms-like tyrosine kinase receptor | sFlt | Reducing angiogenesis |
| Galectin-3 | Gal-3 | Anti-apoptosis |
| Nitric oxide | NO | Vessel dilation |

Other optional serum biomarkers to be studied in a research laboratory include (not limited):
1. senescence markers of p16Ink4a, p21, p53, Density-enhanced phosphatase 1 (DEP1/CD148), and beta-2-microglobulin (B2MG) expression in monocytes
2. telomere length and efferocytosis in monocytes
3. p90RSK activation and ERK5 S496 phosphorylation in monocytes
4. efferocytosis markers (*Gas6*, *Metk, Megf8*), transcription factors (*Pparg/d*, *Klf4*), and inflammatory markers (*Il-1a, Tnfa*, *Mmp12*) in monocytes
5. urine urolithin level
6. p90RSK activation by the response of human monocytes to oxidation

A portion of the serum will be analyzed as described above*.* According to patient consent information, the remaining serum will be stored as de-identified frozen specimens in locked storage in the laboratory of a study team member. As new protocols are developed, they will be presented for IRB review; if approval is obtained, serum may be used for additional research projects.

**4.10 Assessment of cardiac fitness trajectory:**

Patients will be seen before RT, at least twice during RT, and at 3, 6, 12, 18, and 24 months following the completion of RT. At these times, patients will be asked about cardiopulmonary symptoms, and a heart and lung examination will be performed. Heart rate and blood pressure will be recorded. Cardiac events will be graded according to the CTCAE v5. Additionally, patients will complete the Duke Activity Status Index (DASI), a validated assessment of functional capacity that provides an estimate of peak oxygen uptake. Patient reported outcomes using the EQ- 5D-5L, a questionnaire to assess general health status, and MDASI, questionnaire to assess patient symptom burden will be collected as well.

If patients are lost to follow-up, research staff will call them or their family members to obtain follow-up information by phone, as described in the informed consent. Starting at 36 months after RT, a cardiac history will be obtained annually, either in person or by phone, until study termination, disease progression, or patient death, whenever possible.

The procedure for 6-minute walking test is included in Appendix 2.

**4.11 Patient Reported Outcomes (PROs)**

***EQ- 5D-5L:*** The EQ-5D family of instruments has been developed to describe and value health across a wide range of disease areas. They are also frequently used in research into health in the general population. There are three versions of the instrument: EQ-5D-5L, EQ-5D-3L and EQ-5D-Y. For over 25 years, they have been widely used in clinical trials, population studies and in real-world clinical settings. The EQ-5D is used worldwide and has been translated into most major languages through a closely monitored translation process.

Each EQ-5D instrument comprises a short descriptive system questionnaire and a visual analogue scale (EQ VAS) that are cognitively undemanding, taking only a few minutes to complete. The questionnaire provides a simple descriptive profile of a respondent’s health state. The EQ VAS provides an alternative way to elicit an individual’s rating of their own overall current health. When the descriptive system profile is linked to a ‘value set’, a single summary index value for health status is derived that can be used in economic evaluations of healthcare interventions. A value set provides values (weights) for each health state description according to the preferences of the general population of a country/region. Value sets for the EQ-5D-5L and 3L versions are available in a large and growing number of countries Designed for self-completion by respondents and available in both paper and digital versions, the EQ-5D is ideally suited for use in online or postal surveys, in clinics and in interviews (face-to-face or telephone).

The EQ-5D-5L descriptive system comprises the same five dimensions as the EQ-5D-3L (MOBILITY, SELF-CARE, USUAL ACTIVITIES, PAIN / DISCOMFORT and ANXIETY / DEPRESSION), but each dimension now has five response levels: no problems, slight problems, moderate problems, severe problems, unable to /extreme problems. The respondent is asked to indicate his/her health state by checking the box next to the most appropriate response level for each of the five dimensions. Responses are coded as single-digit numbers expressing the severity level selected in each dimension. For instance, ‘slight problems’ (e.g. ‘I have slight problems in walking about’) is always coded as ‘2’. The digits for the five dimensions can be combined in a 5-digit code that describes the respondent’s health state; for instance, 21111 means slight problems in the mobility dimension and no problems in any of the other dimensions The EQ VAS records the respondent’s overall current health on a vertical visual analogue scale, where the endpoints are labelled ‘The best health you can imagine’ and ‘The worst health you can imagine’. The EQ VAS provides a quantitative measure of the patient’s perception of their overall health.

The EQ-5D-5L Questionnaire is included in Appendix 3.

***MDASI-Plus:*** The M. D. Anderson Symptom Inventory (MDASI-Plus) is a valid and reliable tool designed to assess multiple cancer-related symptoms of adults with cancer. This instrument offers several advantages. First, it covers multiple symptoms that commonly occur among adolescent cancer patients, and it is capable of assessing them at one time. It also allows a better understanding of the relationships between multiple symptoms and symptom clusters. We also believe that the rating scale (010 points) is easier and more approachable by adolescents with a wide variety of cognitive abilities, which potentially reduces the patient's burden during assessment. Such an instrument provides a truly usable framework and assessment tool for measuring cancer-related symptoms. This instrument can also be administered electronically.

The MDASI-Plus Questionnaire is included in Appendix 4.

**5.0 Schema, Statistics, and Justification of Sample Size**

**5.1 Schema**

Figure 1 describes the schema for the model development and adaptation.

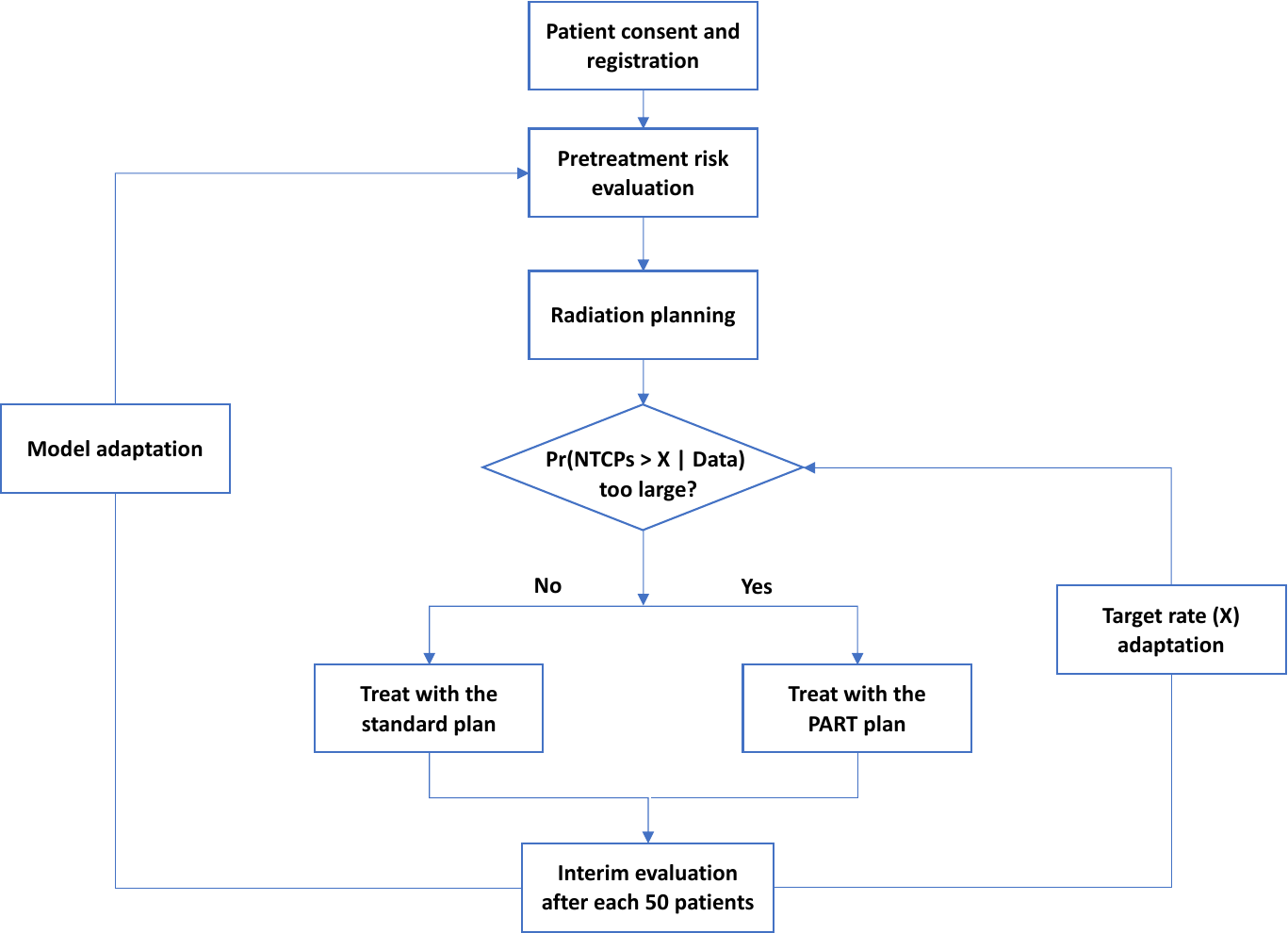

Figure 1: Schema for the model development and adaptation, where X denotes the target NTCP rate we are trying to reduce to.

5.1.1	The standard cardiac dose constraints currently used in practice is shown in Table 3. These dose constraints have been used to calculate the probability of CAEs in NTCP_-S._

| Table 3: Heart dose constraints used in standard and personalized NTCP models | |
| --- | --- |
| NTCP_-S_: NCCN and MDACC | NTCP_-p_: Personalized Heart Dose Constraints |
| MHD≤20Gy | MHD≤20Gy |
| V50≤25%; V45 <=35%; V30 <=50%, | V60≤5%, V75≤2.6%, V80≤0.6% |
|  | V50≤25%; V45 <=35%; V30 <=50% |

5.1.2 We will systematically analyze currently available proton and photon treatment data to develop a “personalized” model of cardiac toxicity that takes into consideration individual patient-specific parameters (e.g., relevant clinical factors, genetic factors, and pre-treatment blood biomarkers), dose-volume indices of individual heart substructures. Examples of clinical factors include age, sex, performance status, tumor histology, tumor location, planning target volume (PTV) and gross tumor volume (GTV) sizes, induction and concurrent chemotherapy, surgery, and comorbidities. Examples of genetic and biomarkers include ASCVD score, CAC, baseline hsTnT and SNP. Dosimetric indices may be in the form or mean dose initially and equivalent uniform dose within each substructure when such data become available. EUD may implicitly consider the impact of high dose subvolumes. Such a model will provide relative contribution of each factor to the predicted heart toxicity and is expected to estimate it more accurately for an individual patient based on his/her specific characteristic for a given treatment plan.
5.1.3 In an analysis of NSCLC patients enrolled to a completed prospective randomized protocol (NCT009105005), we found that the volume of the heart that received 75Gy or higher were associated with the development of grade≥2 cardiac adverse events (CAEs) (V75 = 2.6% with vs.1.4% without CAEs, respectively, p=0.04) for the patients with or without grade >=2 CAEs. Heart V75s were 2.9% and 1.4% for patients with or without grade≥3 CAEs. For this group,16.8% and 12.7% patients developed grade≥2 and grade ≥3 CAEs, respectively. Heart V75≤2.6% were associated with 26.8% and 19.5% grade ≥2 and grade≥3 CAES, respectively.

We also observed that there are 30% patients developed grade>=3 heart toxicity if heart V80 >0.6%. By restricting heart V80 <0.6% in our treatment planning, we can greatly reduce this higher chance of developing grade >=3 heart toxicity.

Heart V60 should be less than 5%. Our IMPT data shows that median OS are 37.7 months and 12.2 months for patient with heart V60 smaller than 5% and larger than 5% with statistical significance (P=0.013). If heart V60<5% dose constraint cannot be met with either plan, we will use the plan with minimized V60<5% for treatment.

For the first NTCP_-P,_ we will use the dose constraints on Table 3 for CAEs prediction, and we expect the X (NTCP_-S_ – NTCP_-P_) ≥ 4%.

5.1.3. For a given patient in the trial, we will use IMPT/IMRT optimization to produce two treatment plans for each modality using
- criteria based on prevalent response model (e.g. dosimetric constraints)
- replacing in the standard criteria the constraints on heart in Table 1 with those listed on table 2 for the heart substructures as predicted by the personalized model above.
- We will calculate NTCP_-S_ and NTCP_-P_ values (see Figure 1) for the heart for both plans using the personalized model described above and compare the two NTCP values.

We will then follow the flow diagram to make treatment decision for each patient in the group until specified number of patients have been accrued.

We will perform interim analyses and modify the model based on the new data and tighten the threshold for difference between NTCPs if indicated.

**5.2 Statistical consideration and sample size calculation**

After the first cohort of 25 patients been enrolled, we developed the first predictive model and we found the accuracy of the model prediction needs to be improved using larger sample size.  As we will conduct model adaptation after each cohort, 25 patients per cohort as originally planned may yield some statistical issues such as non convergence. A larger sample size per cohort is therefore needed to ensure the robustness and reliability of our model adaptation strategy. Also, a larger sample size can yield more accurate estimate of each study endpoint. For example, with the original sample size of 100 patients with 25 patients in each cohort, the maximum width of the 95% confidence interval for an event rate estimate would be 20%. Using a sample size of 200, the maximum width can be reduced to 14%. In addition, the accrual rate to this trial was much faster than originally estimated and we should be able to complete enrollment with expanded samples size without prolong the trial duration. Therefore, the total sample size with be expanded to 200 patients.

We will implement our developed personalized risk prediction model and develop two optimized radiation plans for each enrolled patient: one that uses standard population-based dose constraints used in the standard of practice which will predict NTCP_Standard_ (NTCP_-S_), and the other that uses personalized dose constraints based on individual risk which will predict NTCP_Personlized_ (NTCP_-P_). The trial uses the event of elevation of hs-cTnT and the cardiac event as the co-primary endpoints. Our hypothesis is that the P-model pathway can reduce the probability of either of these two events. Our preliminary results showed 33% incidence at 2 years using standard population-based dose constraints. For this trial, we will consider the incidence of event ≤25% as the goal for both the standard and P-model pathway. In the P-model pathway, if the difference in the predicted normal tissue complication probabilities (DNTCP) (NTCP-_S_-NTCP_- P_) less than certain threshold X% between the two plans, standard treatment will be used; otherwise, a personalized dose will be administered. The initial threshold value X will be determined by the PART model developed in Aim 2. We will use the results based on an analysis of 225 patients who completed a prospective phase 2 trial as the basis to set the goal for the two event rates in the decision making. We plan to conduct several interim analyses during the trial to adaptively validate the PART model with the new information obtained, starting from the completion of cohort 2. The model will be refined as necessary to improve predictive accuracy, and the threshold value X will be adaptively updated based on the data from the concurrent trial. If the P-model pathway shows promising treatment effect in reducing the cardiac events, we will then reduce the threshold magnitude of X to allow more patients to be treated by the personalized dose for the subsequent cohort. The new information obtained from each additional new cohort of patients will help continuous model improvement to minimize cardiotoxicity.

We will simultaneously monitor two co-primary endpoints using the Bayesian optimal phase 2 (BOP2) design^38^. Specifically, let $n$ denote the interim sample size and $N$ denote the maximum sample size. Let $Y_{1}$ and $Y_{2}$ respectively denote the elevation of hs-cTnT and cardiotoxicity, with $Y_{1}=1$ and $Y_{2}=1$ indicating that patients experienced the events in the two respective endpoints. We assume that the joint distribution of $(Y_{1},Y_{2})$ follows a multinomial distribution with 4 elementary outcomes: $(Y_{1},Y_{2})$ = (1, 1), (1, 0), (0, 1) and (0, 0), which indicate the events of (elevation of hs-cTnT, cardiotoxicity), (elevation of hs-cTnT, no cardiotoxicity), (no elevation of hs-cTnT, cardiotoxicity), and (no elevation of hs-cTnT, no cardiotoxicity), respectively.

Let $\mathbf{p}=(P_{11},P_{10},P_{01},P_{00})$ denote the probabilities of observing the four outcomes, and let $p_{1}=Pr(Y_{1}=1)$ (Probability of an elevated hs-cTnT), $p_{2}=Pr(Y_{2}=1)$ (Probability of a cardiotoxicity), and $p_{3}=Pr(Y_{1}=1,Y_{2}=1)$ (Probability of an elevated hs-cTnT and a cardiotoxicity).

The Personalized Plan is deemed as unacceptable if $p_{1}\geq0.25$ and $p_{2}\geq0.2$, i.e., the PART plan is not promising in both co-primary endpoints. The null hypothesis of a 25% hs-cTnT elevation probability and a 20% cardiotoxicity probability is derived based on an analysis of 225 patients who completed a prospective phase 2 trial of CRT for NSCLC. Thus, we will stop enrolling patients and claim the PART plan is not promising if

$$Pr(p_{1}<0.25|data)<\lambda(\frac{n}{N})^{\alpha},$$

**AND**

$$Pr(p_{2}<0.2|data)<\lambda\left( \frac{n}{N} \right)^{\alpha},$$

where $\lambda$=0.98 and $\alpha$=1 are design parameters optimized to maximize the probability of correctly concluding an efficacious treatment as acceptable when $p_{1}=0.15$, $p_{2}=0.15$ and $p_{3}=0.05$, while controlling that the probability of incorrectly claiming an inefficacious treatment, with $p_{1}=0.25$, $p_{2}=0.20$ and $p_{3}=0.10$, as acceptable is less than 5%. This optimization is performed assuming a vague Dirichlet prior $Dir(0.05,0.20,0.15,0.60)$ for $\mathbf{p}$. The prior is chosen such that it corresponds to a prior effective sample size of 1 patient, and the prior estimates of $p_{1}$ and $p_{2}$ match the cutoff values specified above. The above decision rule leads to the following optimal stopping boundaries:

Table 1: Optimized stopping boundaries

| # patients treated | Stop if # elevation >= | **AND** # cardiotoxicity >= |
| --- | --- | --- |
| 50 | 14 | 11 |
| 100 | 24 | 19 |
| 150 | 34 | 26 |
| 200 | 39 | 30 |

Based on Table 1, we perform the interim analysis when the number of enrolled patients reaches 50, 100, 150. When the total number of patients reaches the maximum sample size of 200, we conclude that the treatment is acceptable if the number of toxicities in first endpoint are smaller than 38, or the number of toxicities in second endpoint are smaller than 30; otherwise we conclude that the treatment is unacceptable. The go/no-go criteria in Table 1 are non-binding.

Below are the operating characteristics of the design based on 10000 simulations using the BOP2 web application, which is available at <http://www.trialdesign.org>.

Table 2: Operating characteristics

| Pr(ele) | Pr(tox) | Pr(ele & tox) | Early stopping (%) | Claim promising (%) | Sample size |
| --- | --- | --- | --- | --- | --- |
| 0.25 | 0.20 | 0.10 | 69.91 | 5.03 | 131.8 |
| 0.15 | 0.15 | 0.05 | 0.91 | 96.03 | 199.0 |

##### Note: In scenario 1, the Pr(either an elevation of hs-cTnT or a cardiotoxicity)=40%; in scenario 2, the Pr(either an elevation of hs-cTnT or a cardiotoxicity)=25%. This says that the trial has 95% power to detect the efficacious treatment plan if the treatment effect is 35%-25%=10%, i.e., the PART plan can lower the cardiotoxicity rate by 10%.

The threshold value X will be adaptively updated via a Bayesian posterior learning process. Based on the observed information accrued up to each interim, we will compute the posterior probability that the P-model pathway yields a smaller rate of adverse events than the standard plan among the patients with the DNTCP around the current threshold value X: if this probability is large, we will reduce the threshold value X to allow more patients be treated by P-model pathway; if this probability is particularly small, we will increase the threshold value so that only the high-risk patients will be treated by P-model pathway; otherwise, the threshold value X will remain unchanged. The cutoffs of this posterior probability will be carefully calibrated using simulations.

**5.3 Data analysis plan**

All statistical analyses will be performed by using R software. Patient demographic and baseline characteristics will be summarized using descriptive statistics. Descriptive summary tables will be produced separately for patients treated by the standard plan and the P-model pathway. Using a sample size of 200, the maximum width can be reduced to 14%. Bayesian posterior probability of the PART pathway is better than the historical standard will be calculated, and the Binomial exact test will be performed using a 0.05 significance level. Matched pairs comparison of patients who treated by the PART pathway with historical control patients will be conducted to test the effectiveness of the new treatment plan. As appropriate, we will also assess the relationships between the cardiac event with prognostic factors such as age and gender. As distributions allow, we will also assess the divergence degree between the risk prediction models developed respectively based on the historical data and the data from the new prospective trial.

5.4 Data and Safety Monitoring

Data and Safety Monitoring is the process for reviewing data collected as research progresses to ensure the continued safety of current and future participants as well as the scientific validity and integrity of the research. Studies conducted at MD Anderson will follow the DSMP that has been approved by the NCI.

The Principal Investigator is ultimately responsible for the conduct and monitoring of all aspects of the study on an ongoing basis. The Principal Investigator will provide an annual review and report of the study, including all adverse events, accrual information, efficacy and response data, along with overall study progress and continuation plans to MD Anderson’s Data and Safety Monitoring Committees responsible for study oversight.

**6.0 Data Management and Confidentiality**

**6.1 Data Confidentiality Procedures**

Data will be stored on a password- and firewall-protected computer. Any paper records will be locked in file cabinets in a secured area. The data will be stored until the completion of the final data analysis and publication of study results. They will be destroyed according to MDACC standards at the time of trial termination.

6.2 Data Collection and Management Responsibilities

Data Capture: The REDCap system will be used to manage clinical and laboratory data at MD Anderson. MD Anderson's Research Information Systems and Technology Services (RISTS) group provides deployment and integration support for research data management using the REDCap (Research Electronic Data Capture) application. REDCap allows researchers to create projects with data collection instruments and share those projects with colleagues. Researchers can capture and analyze data through a simple web-based interface. REDCap is a two-tier pHp-based web application that can be hosted on a variety of hardware and operation systems. The application also relies on web server software and a database server to function properly. RISTS works with the Data Center Operations Team (DCOT) to deploy and configure the servers which host REDCap. The application is deployed on a RedHat Linux server that is maintained and monitored by DCOT. The open source Apache web server is used in the REDCap deployment. The servers are backed up nightly and have well-defined processes for disaster recovery. The database used to store project related information is maintained in a MySQL database cluster which is supported by a RISTS Database Administrator (DBA). The database server, like the application server, also has a well-defined process for disaster recovery. The REDCap installation also conforms to the stringent requirements for security and protected health information (PHI). Users are authenticated against MD Anderson’s Active Directory system. External collaborators are given access to projects once approved by the project sponsor. The application is accessed through Secure Socket Layer (SSL).

Clinical data and imaging results will be integrated and managed in REDCap system. Access to project data requires user authentication and is password restricted. There will be no hard copies of data containing patient identifiers. Original subject consent forms will be destroyed after they are scanned and stored on a password protected encrypted server volume within the MD Anderson protected network. Consent forms will not have the subject ID, so there would be no way to use the consent form to re-link the research data to individual patients.

6.3 Confidentiality

All study materials will be identified by study code number and will not have patient identifiers. Study data will be accessible only to study personnel. They will be kept in a locked file cabinet or password-protected computer, with access restricted to study personnel only.

Research data will be kept in a de-identified, password-protected database. A separate database within the RIT, and MD Anderson protected network will contain a key file linking subject ID numbers with patient identifiers. The patient ID database will be kept only as long as needed to complete the data collection and publication. Every effort will be made to protect the patient identifiers, but it is always a possibility that a computer breech could result in loss of the patient ID file. The possibility of this risk is highlighted in the subject consent form.

**7.0 References**

1	American Cancer Society. Lung Cancer Survival Rates. (2020).

2	Lou, Y. *et al.* Survival trends among non-small-cell lung cancer patients over a decade: impact of initial therapy at academic centers. *Cancer Med* **7**, 4932-4942, doi:10.1002/cam4.1749 (2018).

3	Haque, W. *et al.* Trends in Cardiac Mortality in Patients With Locally Advanced Non-Small Cell Lung Cancer. *International Journal of Radiation Oncology Biology Physics* **100**, 470-477, doi:10.1016/j.ijrobp.2017.10.031 (2018).

4	Yegya-Raman, N. *et al.* Dosimetric Predictors of Symptomatic Cardiac Events After Conventional-Dose Chemoradiation Therapy for Inoperable NSCLC. *Journal of Thoracic Oncology* **13**, 1508-1518, doi:10.1016/j.jtho.2018.05.028 (2018).

5	Darby, S. C. *et al.* Risk of ischemic heart disease in women after radiotherapy for breast cancer. *N Engl J Med* **368**, 987-998, doi:10.1056/NEJMoa1209825 (2013).

6	Wang, K. *et al.* Heart dosimetric analysis of three types of cardiac toxicity in patients treated on dose-escalation trials for Stage III non-small-cell lung cancer. *Radiotherapy and Oncology* **125**, 293-300, doi:10.1016/j.radonc.2017.10.001 (2017).

7	Speirs, C. K. *et al.* Heart Dose Is an Independent Dosimetric Predictor of Overall Survival in Locally Advanced Non–Small Cell Lung Cancer. *Journal of Thoracic Oncology* **12**, 293-301, doi:10.1016/j.jtho.2016.09.134 (2017).

8	Liao, Z. *et al.* Bayesian adaptive randomization trial of passive scattering proton therapy and intensity-modulated photon radiotherapy for locally advanced non–small-cell lung cancer. *Journal of Clinical Oncology* **36**, 1813-1822, doi:10.1200/JCO.2017.74.0720 (2018).

9	Elhammali, A. *et al.* Clinical outcomes after intensity-modulated proton therapy with concurrent chemotherapy for inoperable non-small cell lung cancer. *Radiotherapy and Oncology* **136**, 136-142, doi:<https://doi.org/10.1016/j.radonc.2019.03.029> (2019).

10	Shiraishi, Y., Xu, C., Yang, J., Komaki, R. & Lin, S. H. Dosimetric comparison to the heart and cardiac substructure in a large cohort of esophageal cancer patients treated with proton beam therapy or Intensity-modulated radiation therapy. *Radiother Oncol* **125**, 48-54, doi:10.1016/j.radonc.2017.07.034 (2017).

11	Wang, Z. *et al.* Lyman-Kutcher-Burman normal tissue complication probability modeling for radiation-induced esophagitis in non-small cell lung cancer patients receiving proton radiotherapy. *Radiother Oncol* **146**, 200-204, doi:10.1016/j.radonc.2020.03.003 (2020).

12	Palma, G. *et al.* NTCP Models for Severe Radiation Induced Dermatitis After IMRT or Proton Therapy for Thoracic Cancer Patients. *Front Oncol* **10**, 344, doi:10.3389/fonc.2020.00344 (2020).

13	Lai, H. M. *et al.* Association of coronary artery calcium with severity of myocardial ischemia in left anterior descending, left circumflex, and right coronary artery territories. *Clin Cardiol* **35**, 61-63, doi:10.1002/clc.20997 (2012).

14	Kirsch, J. *et al.* Detection of coronary calcium during standard chest computed tomography correlates with multi-detector computed tomography coronary artery calcium score. *Int J Cardiovasc Imaging* **28**, 1249-1256, doi:10.1007/s10554-011-9928-9 (2012).

15	Lakoski, S. G. *et al.* Coronary artery calcium scores and risk for cardiovascular events in women classified as "low risk" based on Framingham risk score: the multi-ethnic study of atherosclerosis (MESA). *Arch Intern Med* **167**, 2437-2442, doi:10.1001/archinte.167.22.2437 (2007).

16	Cardinale, D. *et al.* Trastuzumab-induced cardiotoxicity: clinical and prognostic implications of troponin I evaluation. *J Clin Oncol* **28**, 3910-3916, doi:10.1200/JCO.2009.27.3615 (2010).

17	Sawaya, H. *et al.* Early detection and prediction of cardiotoxicity in chemotherapy-treated patients. *Am J Cardiol* **107**, 1375-1380, doi:10.1016/j.amjcard.2011.01.006 (2011).

18	Adamson, P. D. *et al.* Response to: 'Convalescent troponin and cardiovascular death following acute coronary syndrome' by Kawada. *Heart* **106**, 545-546, doi:10.1136/heartjnl-2020-316547 (2020).

19	Tzolos, E. *et al.* Dynamic Changes in High-Sensitivity Cardiac Troponin I in Response to Anthracycline-Based Chemotherapy. *Clin Oncol (R Coll Radiol)* **32**, 292-297, doi:10.1016/j.clon.2019.11.008 (2020).

20	Pudil, R. Detection of radiation induced cardiotoxicity: Role of echocardiography and biomarkers. *Rep Pract Oncol Radiother* **25**, 327-330, doi:10.1016/j.rpor.2020.02.012 (2020).

21	Onitilo, A. A. *et al.* High-sensitivity C-reactive protein (hs-CRP) as a biomarker for trastuzumab-induced cardiotoxicity in HER2-positive early-stage breast cancer: a pilot study. *Breast Cancer Res Treat* **134**, 291-298, doi:10.1007/s10549-012-2039-z (2012).

22	Canada, J. M. *et al.* Increased C-reactive protein is associated with the severity of thoracic radiotherapy-induced cardiomyopathy. *Cardiooncology* **6**, 2, doi:10.1186/s40959-020-0058-1 (2020).

23	Wen, J. *et al.* Genetic variants of the LIN28B gene predict severe radiation pneumonitis in patients with non-small cell lung cancer treated with definitive radiation therapy. *Eur J Cancer* **50**, 1706-1716, doi:10.1016/j.ejca.2014.03.008 (2014).

24	Xiong, H. *et al.* ATM Polymorphisms Predict Severe Radiation Pneumonitis in Patients With Non-Small Cell Lung Cancer Treated With Definitive Radiation Therapy. *International Journal of Radiation Oncology*Biology*Physics* **85**, 1066-1073, doi:<https://doi.org/10.1016/j.ijrobp.2012.09.024> (2013).

25	Schlaak, R. A. *et al.* Differences in Expression of Mitochondrial Complexes Due to Genetic Variants May Alter Sensitivity to Radiation-Induced Cardiac Dysfunction. *Front Cardiovasc Med* **7**, 23, doi:10.3389/fcvm.2020.00023 (2020).

26	Walker, V. *et al.* Early detection of subclinical left ventricular dysfunction after breast cancer radiation therapy using speckle-tracking echocardiography: association between cardiac exposure and longitudinal strain reduction (BACCARAT study). *Radiat Oncol* **14**, 204, doi:10.1186/s13014-019-1408-8 (2019).

27	Jones, L. W. *et al.* Prognostic significance of functional capacity and exercise behavior in patients with metastatic non-small cell lung cancer. *Lung Cancer* **76**, 248-252, doi:10.1016/j.lungcan.2011.10.009 (2012).

28	Schmidt, K., Vogt, L., Thiel, C., Jäger, E. & Banzer, W. Validity of the six-minute walk test in cancer patients. *Int J Sports Med* **34**, 631-636, doi:10.1055/s-0032-1323746 (2013).

29	Granger, C. L., Holland, A. E., Gordon, I. R. & Denehy, L. Minimal important difference of the 6-minute walk distance in lung cancer. *Chron Respir Dis* **12**, 146-154, doi:10.1177/1479972315575715 (2015).

30	Miller, K. L. *et al.* Preliminary report of the 6-minute walk test as a predictor of radiation-induced pulmonary toxicity. *Int J Radiat Oncol Biol Phys* **62**, 1009-1013, doi:10.1016/j.ijrobp.2004.12.054 (2005).

31	Siegel, R. L., Miller, K. D. & Jemal, A. Cancer statistics, 2020. *CA: A Cancer Journal for Clinicians* **70**, 7-30, doi:10.3322/caac.21590 (2020).

32	Ray, K. K. *et al.* The ACC/AHA 2013 guideline on the treatment of blood cholesterol to reduce atherosclerotic cardiovascular disease risk in adults: the good the bad and the uncertain: a comparison with ESC/EAS guidelines for the management of dyslipidaemias 2011. *European Heart Journal* **35**, 960-968, doi:10.1093/eurheartj/ehu107 (2014).

33	Gayed, I. *et al.* The clinical implications of myocardial perfusion abnormalities in patients with esophageal or lung cancer after chemoradiation therapy. *Int J Cardiovasc Imaging* **25**, 487-495, doi:10.1007/s10554-009-9440-7 (2009).

34	Marks, L. B. *et al.* The incidence and functional consequences of RT-associated cardiac perfusion defects. *Int J Radiat Oncol Biol Phys* **63**, 214-223, doi:10.1016/j.ijrobp.2005.01.029 (2005).

35	Cheitlin, M. D. *et al.* ACC/AHA/ASE 2003 guideline update for the clinical application of echocardiography: summary article. *Journal of the American College of Cardiology* **42**, 954-970, doi:doi:10.1016/S0735-1097(03)01065-9 (2003).

36	Lancellotti, P. *et al.* Expert consensus for multi-modality imaging evaluation of cardiovascular complications of radiotherapy in adults: a report from the European Association of Cardiovascular Imaging and the American Society of Echocardiography. *European Heart Journal - Cardiovascular Imaging* **14**, 721-740, doi:10.1093/ehjci/jet123 (2013).

37	Armenian, S. H. *et al.* Prevention and Monitoring of Cardiac Dysfunction in Survivors of Adult Cancers: American Society of Clinical Oncology Clinical Practice Guideline. *Journal of Clinical Oncology* **35**, 893-911, doi:10.1200/jco.2016.70.5400 (2017).

38	Zhou, H., Lee, J. J., Yuan, Y. BOP2: Bayesian optimal design for phase II clinical trials with simple and complex endpoints. *Statistics in medicine* **36**, 3302-3314, doi: <https://doi.org/10.1002/sim.7338> (2017).
